# Changes in UK hospital mortality in the first wave of COVID-19: the ISARIC WHO Clinical Characterisation Protocol prospective multicentre observational cohort study

**DOI:** 10.1101/2020.12.19.20248559

**Authors:** Annemarie B Docherty, Rachel H Mulholland, Nazir I Lone, Christopher P Cheyne, Daniela De Angelis, Karla Diaz-Ordaz, Cara Donoghue, Thomas M Drake, Jake Dunning, Sebastian Funk, Marta García-Fiñana, Michelle Girvan, Hayley E Hardwick, Janet Harrison, Antonia Ho, David M Hughes, Ruth H Keogh, Peter D Kirwan, Gary Leeming, Jonathan S Nguyen-Van-Tam, Riinu Pius, Clark D Russell, Rebecca Spencer, Brian DM Tom, Lance Turtle, Peter JM Openshaw, J Kenneth Baillie, Ewen M Harrison, Malcolm G Semple, for ISARIC4C investigators

**Author notes:** Corresponding author: Annemarie B Docherty, Senior Clinical Lecturer, Centre for Medical Informatics, Usher Institute, University of Edinburgh, Edinburgh EH16 4UX, 07916 170395, @abdocherty79. joint first authors (Annemarie B Docherty, Rachel H Mulholland). joint last authors (Ewen M Harrison, Malcolm G Semple). Annemarie Docherty, University of Edinburgh, Senior Clinical Lecturer and Honorary Consultant in Critical Care. Rachel H Mulholland, University of Edinburgh, BREATHE Hub Data Analyst. Nazir I Lone, University of Edinburgh, Senior Clinical Lecturer and Honorary Consultant in Critical Care. Christopher P Cheyne, Department of Health Data Science, Institute of Population Health, University of Liverpool, Research Associate. Daniela De Angelis, University of Cambridge, Professor of Statistical Science for Health. Karla Diaz-Ordaz, Department of Medical Statistics, London School of Hygiene and Tropical Medicine, UK, Associate Professor. Cara Donoghue, University of Liverpool, Project and Data Administrator. Thomas M Drake, University of Edinburgh, Research Fellow. Jake Dunning, Public Health England, Head of Emerging Infections and Zoonoses. Sebastian Funk, Department of Medical Statistics, London School of Hygiene and Tropical Medicine, UK. Marta Garcia-Finana, Department of Health Data Science, Institute of Population Health, University of Liverpool, Professor of Biostatistics. Michelle Girvan, University of Liverpool, Data Manager. Hayley E Hardwick, University of Liverpool, Project Manager. Janet Harrison, University of Liverpool, Data Manager. Antonia Ho, University of Glasgow, Clinical Senior Lecturer and Honorary Consultant in Infectious Diseases. David M Hughes, Department of Health Data Science, Institute of Population Health, University of Liverpool, Lecturer. Ruth H Keogh, Department of Medical Statistics, London School of Hygiene and Tropical Medicine, UK, Professor of Biostatistics & Epidemiology. Peter D Kirwan, MRC Biostatistics Unit, University of Cambridge ROLE? Gary Leeming, University of Liverpool, Senior Data Manager. Jonathan S Nguyen-Van-Tam, University of Nottingham School of Medicine, Professor of Health Protection. Riinu Pius, University of Edinburgh, Senior Data Manager. Clark D Russell, University of Edinburgh, Clinical Lecturer. Rebecca Spencer, University of Liverpool, Project and Data Administrator. Brian DM Tom, MRC Biostatistics Unit, University of Cambridge, Lecturer in Statistics. Lance Turtle, University of Liverpool, Senior Clinical Lecturer in Infectious Diseases. Peter JM Openshaw, Imperial College London, Professor of Experimental Medicine. J Kenneth Baillie, University of Edinburgh, Academic Consultant in Critical Care Medicine. Ewen M Harrison, University of Edinburgh, Professor of Surgery and Data Science and Honorary Consultant Surgeon. Malcolm G Semple, University of Liverpool Professor of Outbreak Medicine and Child Health, Alder Hey Children’s NHS Foundation Trust Liverpool Consultant Physician in Paediatric Respiratory Medicine.

## Abstract

**Background:** Mortality rates of UK patients hospitalised with COVID-19 appeared to fall during the first wave. We quantify potential drivers of this change and identify groups of patients who remain at high risk of dying in hospital.

**Methods:** The International Severe Acute Respiratory and emerging Infection Consortium (ISARIC) WHO Clinical Characterisation Protocol UK recruited a prospective cohort admitted to 247 acute UK hospitals with COVID-19 in the first wave (March to August 2020). Outcome was hospital mortality within 28 days of admission. We performed a three-way decomposition mediation analysis using natural effects models to explore associations between week of admission and hospital mortality adjusting for confounders (demographics, comorbidity, illness severity) and quantifying potential mediators (respiratory support and steroids).

**Findings:** Unadjusted hospital mortality fell from 32.3% (95%CI 31.8, 32.7) in March/April to 16.4% (95%CI 15.0, 17.8) in June/July 2020. Reductions were seen in all ages, ethnicities, both sexes, and in comorbid and non-comorbid patients. After adjustment, there was a 19% reduction in the odds of mortality per 4 week period (OR 0.81, 95%CI 0.79, 0.83). 15.2% of this reduction was explained by greater disease severity and comorbidity earlier in the epidemic. The use of respiratory support changed with greater use of non-invasive ventilation (NIV). 22.2% (OR 0.94, 95%CI 0.94, 0.96) of the reduction in mortality was mediated by changes in respiratory support.

**Interpretation:** The fall in hospital mortality in COVID-19 patients during the first wave in the UK was partly accounted for by changes in case mix and illness severity. A significant reduction was associated with differences in respiratory support and critical care use, which may partly reflect improved clinical decision making. The remaining improvement in mortality is not explained by these factors, and may relate to community behaviour on inoculum dose and hospital capacity strain.

**Funding:** NIHR & MRC

**Key points / Research in Context:** *Evidence before this study:* Risk factors for mortality in patients hospitalised with COVID-19 have been established. However there is little literature regarding how mortality is changing over time, and potential explanations for why this might be. Understanding changes in mortality rates over time will help policy makers identify evolving risk, strategies to manage this and broader decisions about public health interventions.

*Added value of this study:* Mortality in hospitalised patients at the beginning of the first wave was extremely high. Patients who were admitted to hospital in March and early April were significantly more unwell at presentation than patients who were admitted in later months. Mortality fell in all ages, ethnic groups, both sexes and in patients with and without comorbidity, over and above contributions from falling illness severity. After adjustment for these variables, a fifth of the fall in mortality was explained by changes in the use of respiratory support and steroid treatment, along with associated changes in clinical decision-making relating to supportive interventions. However, mortality was persistently high in patients who required invasive mechanical ventilation, and in those patients who received non-invasive ventilation outside of critical care.

*Implications of all the available evidence:* The observed reduction in hospital mortality was greater than expected based on the changes seen in both case mix and illness severity. Some of this fall can be explained by changes in respiratory care, including clinical learning. In addition, introduction of community policies including wearing of masks, social distancing, shielding of vulnerable patients and the UK lockdown potentially resulted in people being exposed to less virus. The decrease in mortality varied depending on the level of respiratory support received. Patients receiving invasive mechanical ventilation have persistently high mortality rates, albeit with a changing case-mix, and further research should target this group. Severe COVID-19 disease has primarily affected older people in the UK. Many of these people, but not all have significant frailty. It is essential to ensure that patients and their families remain at the centre of decision-making, and we continue with an individualised approach to their treatment and care.

## Introduction

There is growing evidence that mortality from COVID-19 is falling, both in hospital and in the community.^1-4^ One explanation may be that the case-mix of patients presenting to hospital has changed towards a younger and less comorbid demographic, who were at lower risk of dying. National UK lockdown and effective shielding measures of vulnerable at-risk populations may have reduced transmission of the virus. Easier accessibility to testing as well as advice regarding seeking medical help may have resulted in earlier presentation to hospital. Familiarity with the virus and clinical course may have led to better management of patients through improved ward and ICU care.^5,6^ Corticosteroids have been shown in trials to reduce mortality in patients with severe COVID-19.^7,8^

The International Severe Acute Respiratory and emerging Infections Consortium (ISARIC) WHO Clinical Characterisation Protocol UK (CCP-UK)^9,10^ was activated on 17 January 2020 to recruit COVID-19 patients admitted to a network of hospitals in England, Scotland and Wales.^11^ ISARIC has been prepared for outbreaks such as COVID-19 for the past 8 years with the intent that it provide data and samples for near real-time analysis.^9,10^ During the COVID-19 outbreak, analysis of CCP-UK cohort in the first wave allowed development of the pragmatic ISARIC 4C Mortality Score for hospitalised COVID-19 patients in readiness to aid management decisions in wave 2.^12^

We aimed to use the ISARIC CCP-UK cohort to describe how 28-day in-hospital mortality has changed over time in patients hospitalised with COVID-19. We explored potential drivers for these changes by assessing the patient characteristics, the severity of illness and the treatment they received during their hospital admission.

## Methods

### Study design and setting

We undertook a prospective cohort study using participants in the ISARIC WHO CCP-UK cohort who were admitted to acute general hospitals (n=247) in the first wave: 9 March to 2 August 2020. National strategy changed from containment to admission based on clinical need on 12 March.^13^

### Participants and study size

Recruited patients were adults (≥18yrs) admitted to hospital with high likelihood or confirmed COVID-19 from assumed community acquired infection (further details in online supplement). All patients were admitted at least six weeks prior to data extraction to allow for sufficient 28 day follow up. For the main analysis, we excluded patients without an outcome date (included as “survivors” in online supplement sensitivity analysis).

### Data collection

Data were extracted from routine healthcare records and recorded onto case report forms on a REDCap database. Under the Control of Patient Information (COPI) notice 2020 for urgent public health research, processing of demographic and routine clinical data from medical records for research does not require consent in England and Wales.^14^ In Scotland, a waiver for consent was obtained from the Public Benefit and Privacy Panel.^15^

#### Exposures

The main exposure of interest was the week of admission to hospital, defined using the ISO week date (ordinal week of the year). To facilitate comparison across time periods, this was also categorised into 3 equal time-periods (TP): TP1 = ISO weeks 11 to 17 (9 March to 26 April), TP2 = 18 to 24 (27 April to 14 June) and TP3 = 25 to 31 (15 June to 2 August).

#### Variables

We collected information on key variables including patient characteristics, illness severity, level of respiratory support, COVID-19 specific treatments, and hospital mortality.

Patient characteristics: patient age (<50yrs, 50-69yrs, 70-79yrs, 80yrs+); sex (Female, Male); self-reported ethnicity (South Asian, East Asian, Black, Other Ethnic Minority and White)^16^, index of multiple deprivations (derived from individual patient postal code), and health worker. Modified Charlson comorbidities (see online supplement) were used to construct the comorbidity count (0, 1, 2+).

Severity of illness at admission (or within 24h) was assessed using physiological components of the ISARIC 4C Mortality Score^12^ : respiratory rate (RR, breaths/min), peripheral oxygen saturation on room air (SpO2, %), Glasgow coma scale (GCS), urea (mmol/L) and C-reactive protein (CRP, mg/L). To capture the patterns in accessing hospital treatment we calculated the days from symptom onset to admission.

Patients were categorised as managed on the ward or in critical care (see the online supplement for more information on level of respiratory support). Maximum level of respiratory support was categorised into: no respiratory support, oxygen (face mask/nasal cannulae/high flow nasal oxygen (HFNO)), non-invasive ventilation (NIV), and invasive mechanical ventilation (IMV).

COVID-19 specific treatments: we recorded only whether patients received corticosteroids (dexamethasone, hydrocortisone, methylprednisolone and prednisolone), as these were the only treatment with proven mortality benefit for COVID-19 in randomised controlled trials.^7,17^

#### Outcomes

The primary outcome was the weekly in-hospital 28-day mortality, as a proportion of all patients admitted in the observed week. Mortality was defined as an outcome of death or discharge to palliative care. The 28-day threshold aligns with the Public Health England definition of a COVID-19 death.^18^

Secondary outcomes were changes in patient demographics and illness severity for patients managed on the ward and in critical care. Within critical care, we looked separately at oxygen only, NIV, and IMV. Within ward care, we looked at those receiving no respiratory support, oxygen only and NIV.

### Statistical methods

Continuous data are presented as mean (standard deviation) or median (interquartile range) depending on the distribution. Categorical data are presented as frequency (%). For univariable comparisons, we used Welch’s *t*-test, ANOVA, Mann–Whitney U, or Kruskal–Wallis tests according to data distribution. Categorical data were compared using chi-squared tests. Counts and proportions for each of the exposure variables were calculated across the three equal time-periods. The weekly proportion of patients admitted and in-hospital 28-day mortality were stratified by each explanatory variable of interest. 95% confidence intervals for these proportions were calculated using the exact method.

There were missing data due to the challenges of real-time data collection during a pandemic. Missing data are reported, and patterns of missing data were explored. Missing values for comorbidities, healthcare worker status, and respiratory support (receiving oxygen, invasive and non-invasive ventilation) were classified as ‘No’. For the primary analysis, multiple imputation using chained equations was performed for missing markers of illness severity. Ten sets, each with 10 iterations, were imputed using 35 explanatory variables including outcomes. Graphical checks of convergence were performed. All analyses were performed using imputed datasets.

#### Primary outcome

Our modelling strategy was informed by a putative causal model (Figure E1 proposed directed acyclic graph). Using logistic regression, we specified three models exploring the association between admission week (as a continuous variable) and hospital mortality. A baseline model included adjustment for age and sex. A second model accounted for known baseline confounders including variables previously shown to be associated with in-hospital mortality: age, sex, comorbidity count, index of multiple deprivation and severity of illness (RR, SpO2, GCS, urea, CRP). In a third model, potential mediators were added to explore the effect of treatment (steroids and respiratory support, including clinical decision making for the latter) on the association between admission week and mortality (controlled direct effect models). We extended this in a potential outcomes framework to perform a three-way decomposition mediation analysis using natural effects models.^19^ We sought to control confounding between, i) exposure and outcome, ii) exposure and mediator, iii) mediator and outcome, iv) and we considered carefully potential mediator:outcome confounders influenced by the exposure. Using standard frequentist approaches, unobserved nested counterfactuals were imputed using an outcome model to accommodate our nominal mediator (respiratory support). Exposure:mediator interactions were explored and a joint model used to incorporate steroid use. Robust standard errors (based on sandwich estimator) were generated, and results presented as a proportion mediated on the risk difference scale.

#### Secondary outcome

To better understand patterns of mortality for different levels of respiratory support, time series data were modelled using Bayesian generalised additive models (GAMs) to allow easy incorporation of multiply imputed datasets (see online supplement for more details).

All analyses were conducted using R (version 3.6.3) and Stan using the packages tidyverse, finalfit, brms, mgcv, mice, medflex, gridExtra and cowplot.

## Results

### Patient inclusion

The final cohort contained 63,972 patients (Table 1, Figure E2), from 247 acute hospitals in England, Scotland and Wales, approximately 48% of all hospital admissions.^1^ Admissions peaked in late March and in early April for all age groups, and steadily decreased until the end of the study (Figure 1A). 29% (N=15,864) of patients managed on the ward died within 28 days of admission, compared with 36% (N=3,317) within critical care (Figure E2).

**Table 1:**
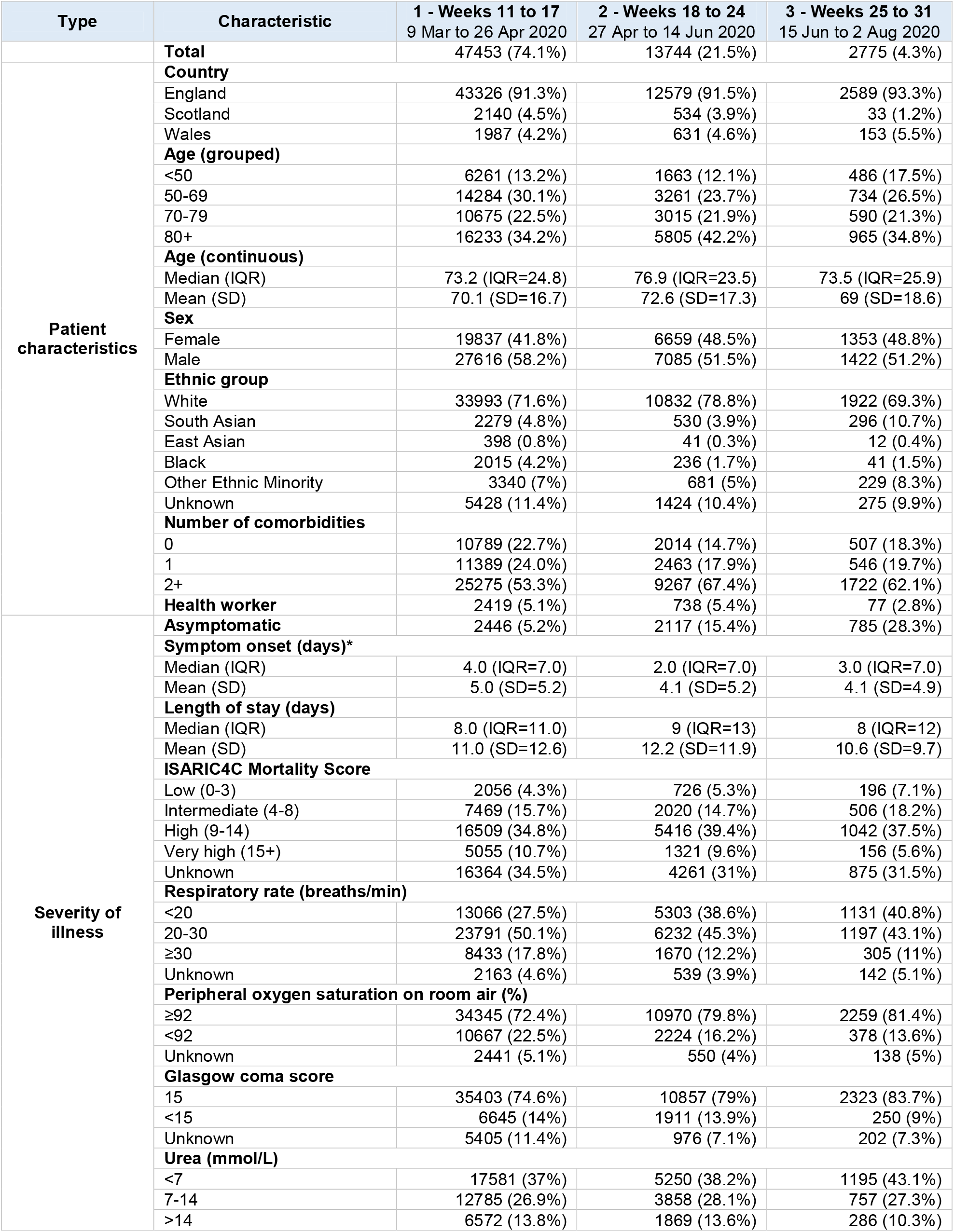
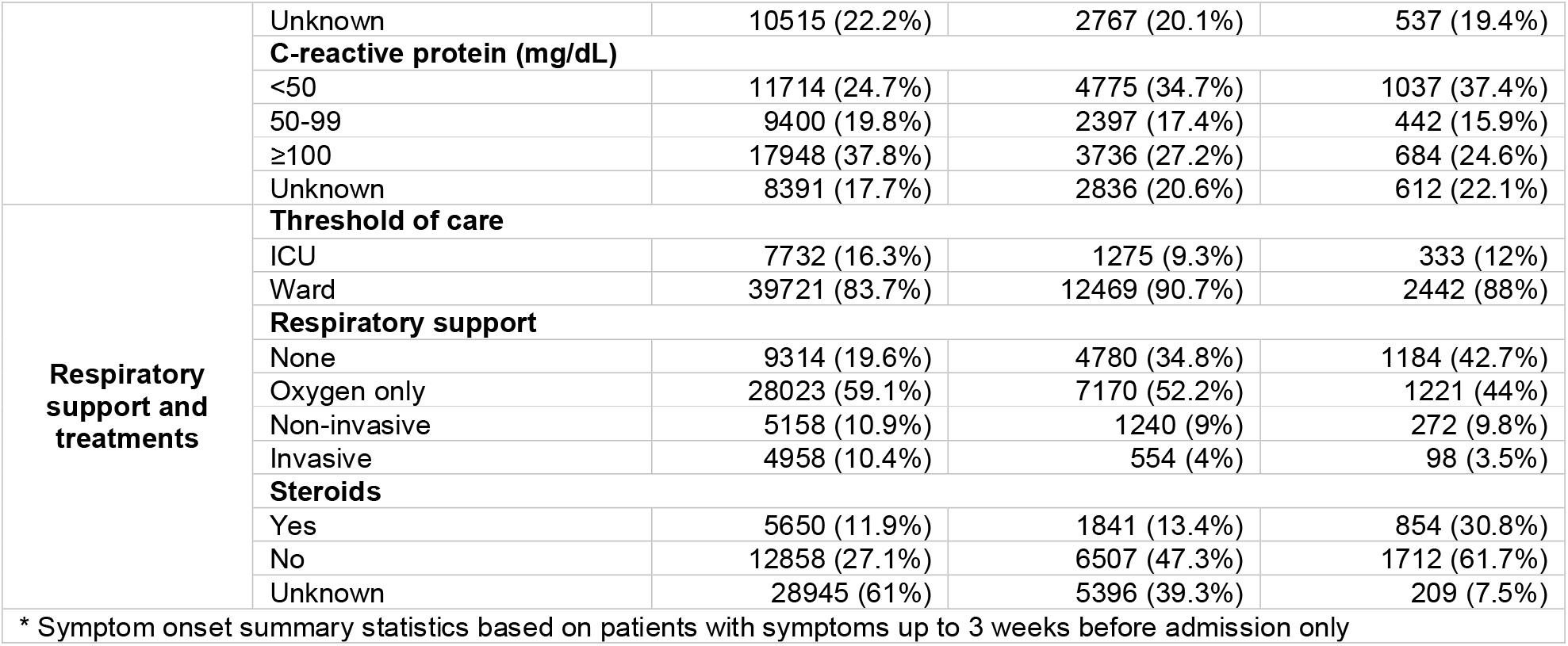
Baseline characteristics of adult patients admitted to hospital with COVID-19, stratified by time (N=63,972).

**Figure 1:**
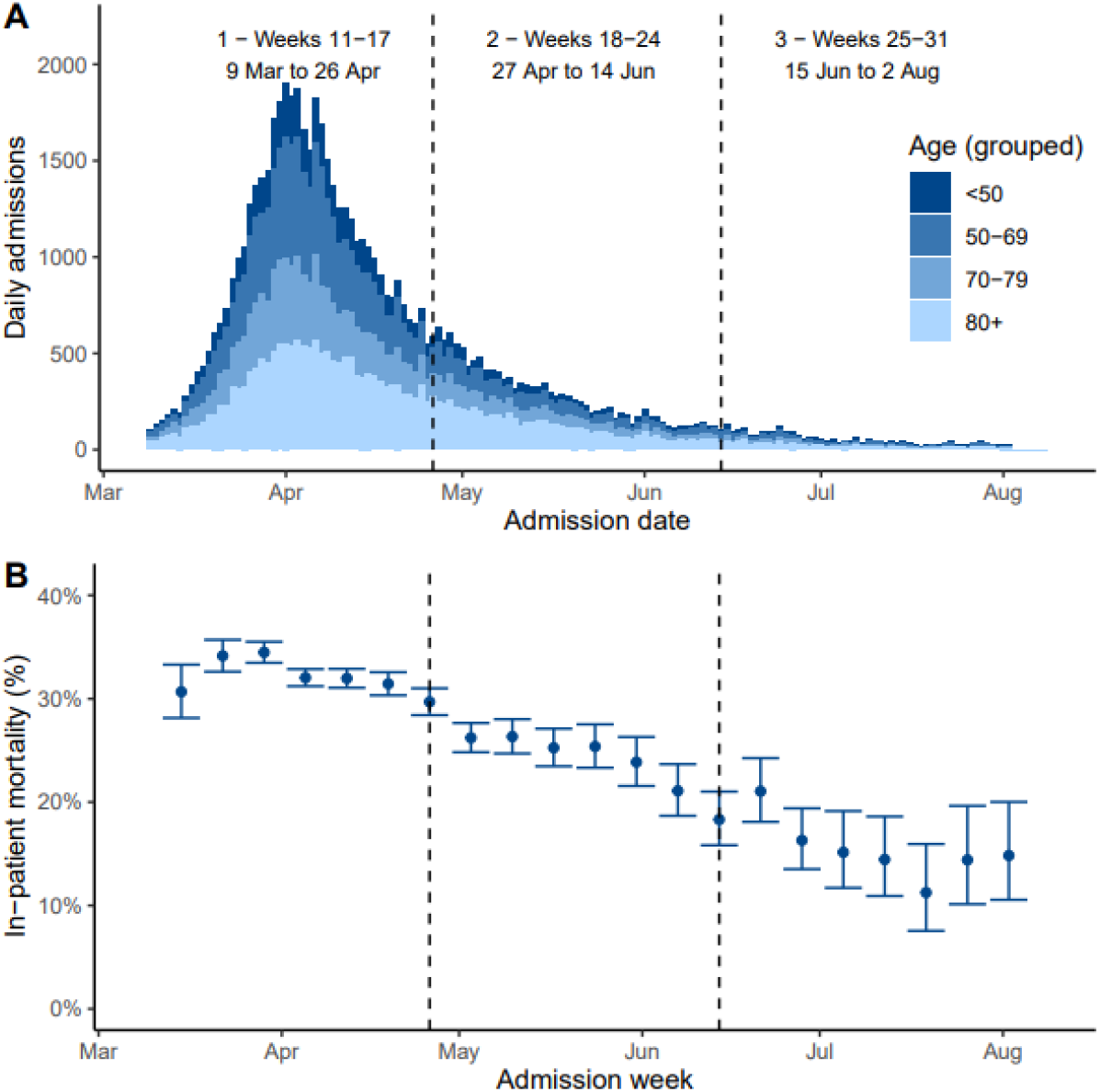
(A) Daily adult COVID-19 admissions from 9 March to 2 August 2020 by age. (B) Weekly unadjusted in-hospital mortality and 95% confidence intervals for adults admitted with COVID-19 from 9 March 2020 to 2 August 2020. 95% confidence intervals calculated by the Exact method. Divided into 3 equal time periods (Weeks 11 to 17, 18 to 24 and 25 to 31).

#### Patient demographics and severity of illness

The majority of patients admitted throughout the first wave were ≥50yrs (N=55,562 (87%)). There was an increase in the proportion of younger people (<50yrs) admitted over time (TP1 13.2% vs TP3 17.5%) (Table 1, Figure 2A). There were initially more men admitted than women (60%:40%), but proportions were similar from mid-April (Figure 2B). The population was multi-morbid, with over 50% of patients having two or more comorbidities and this increased over time (Figure 2C, Table E1). Patients were mostly of White ethnicity with an increasing proportion of South Asian and a decreasing proportion of Black ethnic groups (Figure 2D). The most deprived quintile was most prevalent in all time periods and increased over time (Figure 2E).

**Figure 2:**
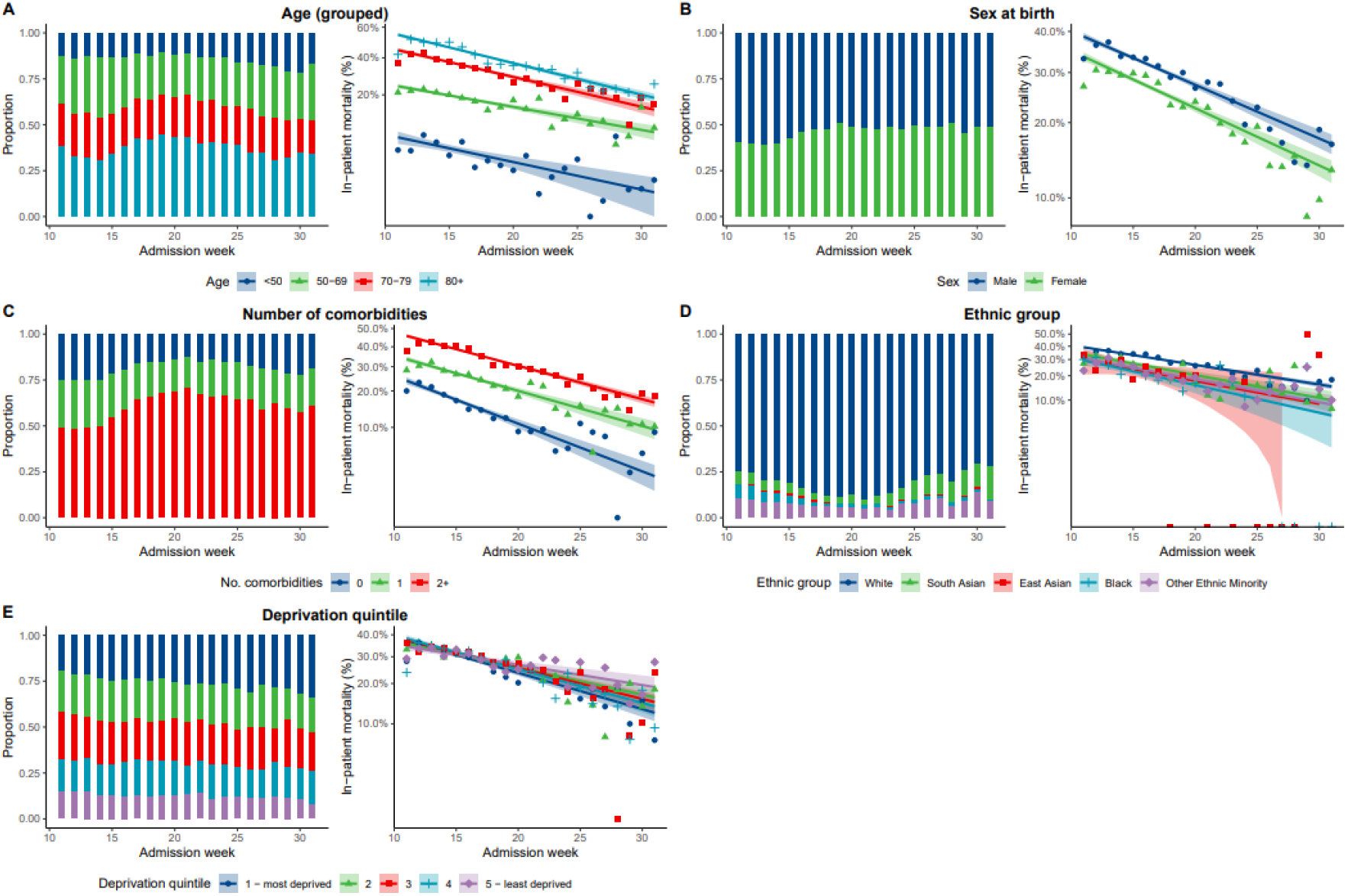
(Left) Adults admitted to hospital with COVID-19 by time stratified by patient characteristics (%). (Right) Unadjusted in-hospital mortality by time stratified by patient characteristics (%). Unknown measurements are excluded from this figure.

Illness severity peaked around March 30 to April 12, when people at presentation to hospital had faster respiratory rates, lower peripheral oxygen saturations, and higher rates of reduced conscious level, acute kidney injury and inflammation, compared with patients admitted subsequently (Figure 3). Patients presented later in their disease process at the beginning compared with the end of the first wave (TP1 med 4 days (IQR 7), TP2 med 2 (IQR 7), TP3 med 3 (IQR 7)) (Table 1).

**Figure 3:**
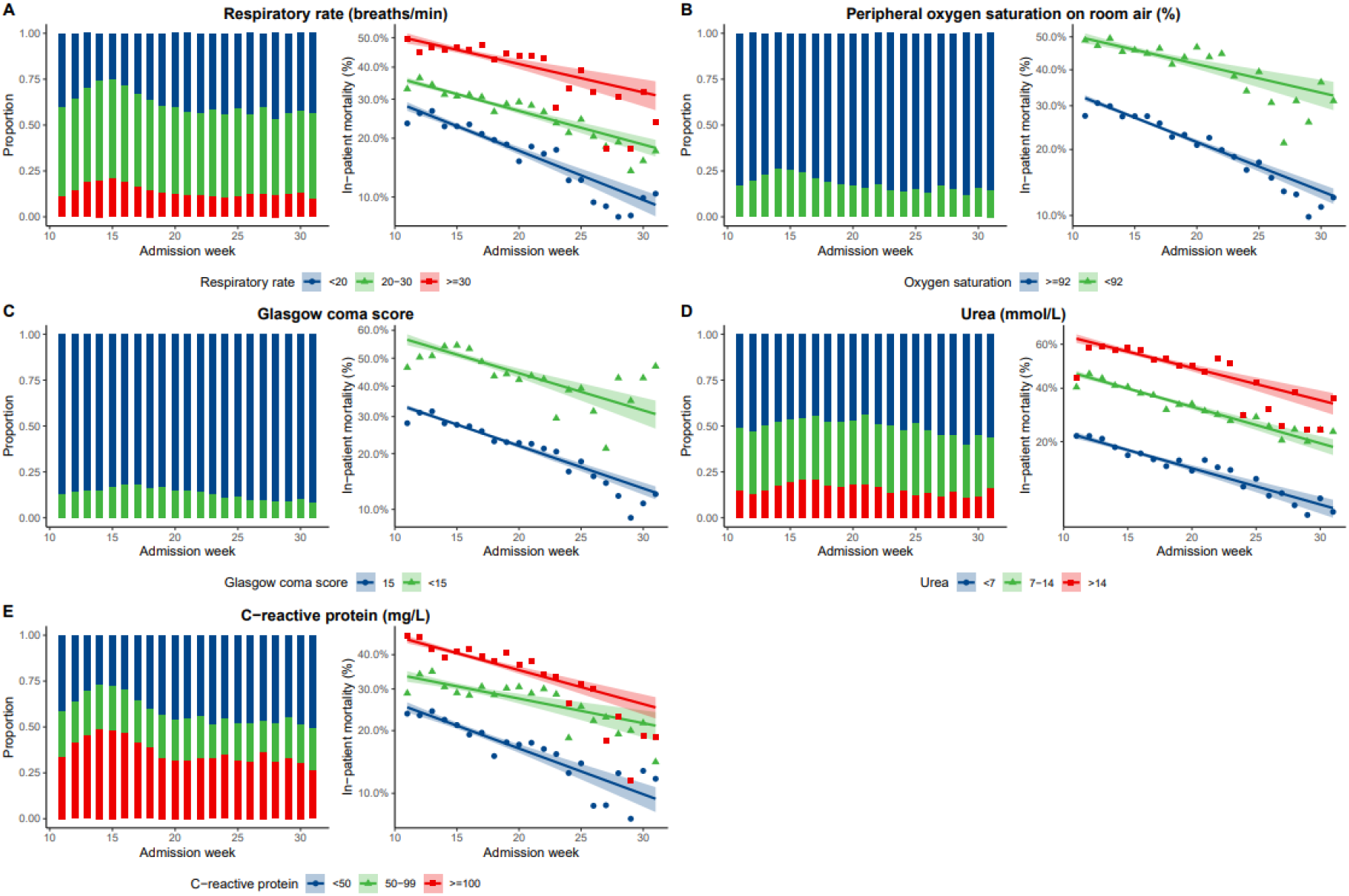
(Left) Proportion of markers of severity of illness at admission to hospital by week of admission. (Right) Unadjusted in-hospital mortality rate per category by week of admission. Unknown measurements are excluded from this figure.

#### Respiratory support and critical care admission

At the peak of admissions, over 80% of patients admitted to hospital received supplementary oxygen. This reduced consistently over subsequent weeks to around 50% for patients admitted in July onwards (Figure E3).

Most patients were managed on the ward, with the proportion of patients being admitted to critical care peaking at the start of the study (N=7,732 (16.3%)) (Table 1, Figure E4). Patients admitted to critical care were younger, and were more likely to be male (Critical care male N=6,433 (68.9%) vs ward male N=29,690 (54.3%)) (Tables E2, E3). Patients with multi-morbidity accounted for a significant proportion of patients (Critical care comorbid N= 3,733 (40%) vs ward comorbid N= 32,531 (60%)) (Tables E2, E3). The pattern of increasing proportions over time of younger and more comorbid patients in critical care mirrored that seen on the ward (Tables E2, E3).

Level of respiratory support received reduced over time for both critical care and ward patients (Figure 4). As the requirement for IMV fell (TP1 64%, TP3 29%, Table E2), the proportion of those requiring NIV substantially increased from 23% to 47% (Table E2). In comparison, on the ward, NIV proportions remained very low decreasing from 9% to 5% (Table E3). By the end of the first wave, 42.7% of all patients admitted received no respiratory support (ward patients 48.5%) (Table 1, Table E3, Figure 4). More information on patient characteristics within Critical care and the ward and respiratory support groups are in the online supplement (Tables E4-E9).

**Figure 4:**
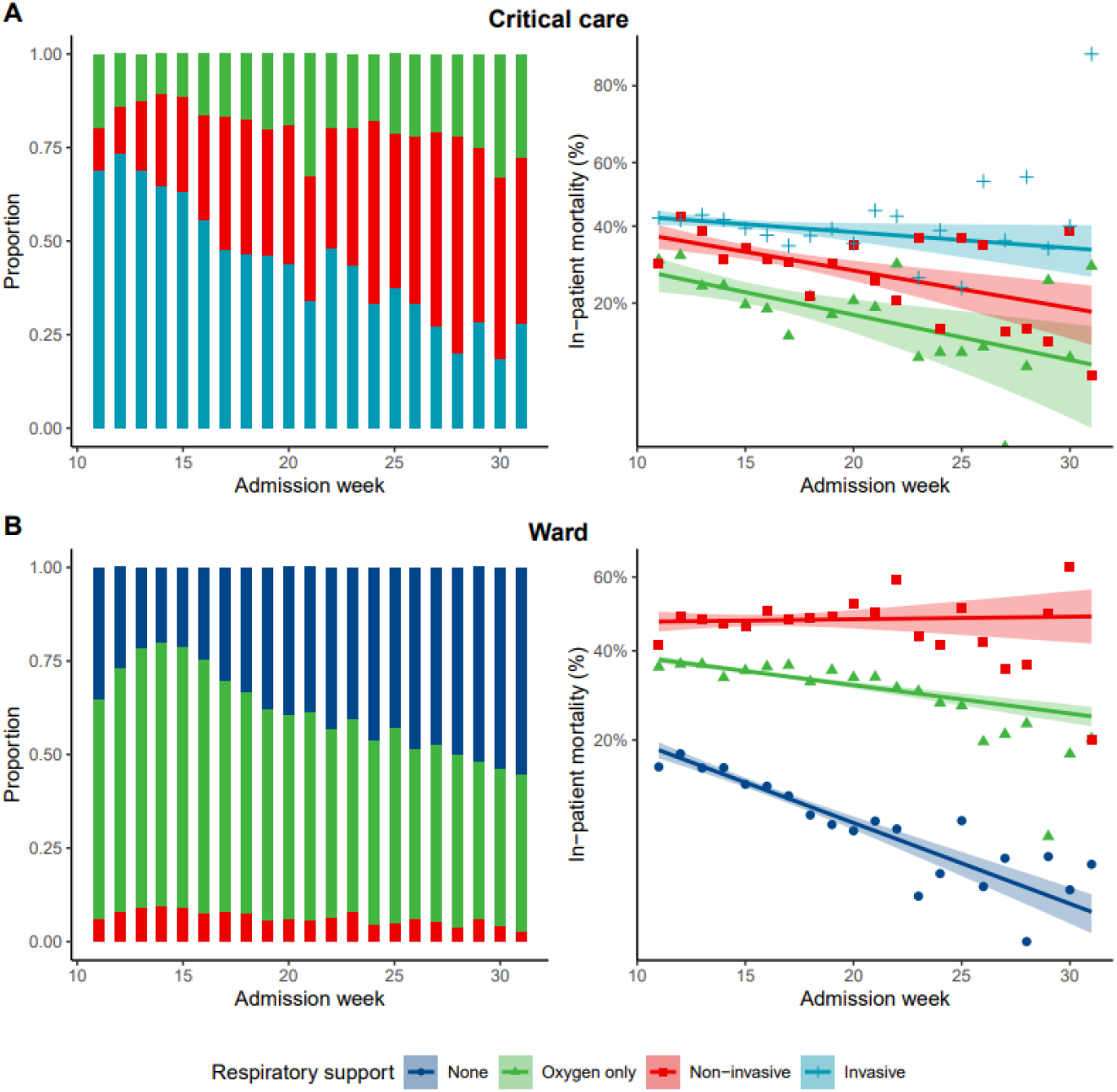
Respiratory support within Critical Care (top) and ward (bottom). (Left) Proportion of respiratory support treatments by week of admission. (Right) Unadjusted in-hospital mortality rate per category by week of admission.

#### Steroid use

The proportion of patients who received steroids increased from 12% (22% in critical care) at the start of the pandemic to 31% (65% in critical care) in June and July (Tables 1 and E2, Figure E5 & E6), mainly in the groups receiving respiratory support.

#### Hospital mortality

In-hospital mortality in patients admitted in March and early April fell from 32% to 16% for patients admitted in June and July. (Figure 1B, Table E10). This did not substantially differ in the sensitivity analysis including those without an outcome reclassified as ‘survivors’ (Figure E7).

Hospital mortality was higher with increasing age, increasing comorbidity count, and male sex, and fell for all demographic categories, most notably in the older (80+ yrs TP1 mortality 48.0% vs TP3 25.0%) and comorbid populations (Figure 2, Table E10). Markers of increased severity of illness at presentation to hospital were associated with increased in-hospital mortality. Mortality fell for all markers of severity of illness over time (Figure 3) and for patients treated in both ward and ICU environments (Figure E4).

There was a 22% reduction in odds of mortality per 4 week period (OR 0.78, 95%CI 0.77 to 0.80, Figure 6). After adjustment for age and sex, this was OR 0.74 (95% CI 0.72, 0.75). After additional adjustment for severity and comorbidity, the effect of week of admission on hospital mortality was similar (OR 0.77, 95% CI 0.75 to 0.78). With addition of the mediator variables (respiratory support/steroids), the OR reduced to 0.81 (95% CI 0.79 to 0.83). 22.2% (OR 0.95, 95% CI 0.94 to 0.96) of the effect of week of admission on hospital mortality was mediated through respiratory care and steroids. There was a significant interaction between respiratory support and week of admission (p<0.001).

**Figure 5:**
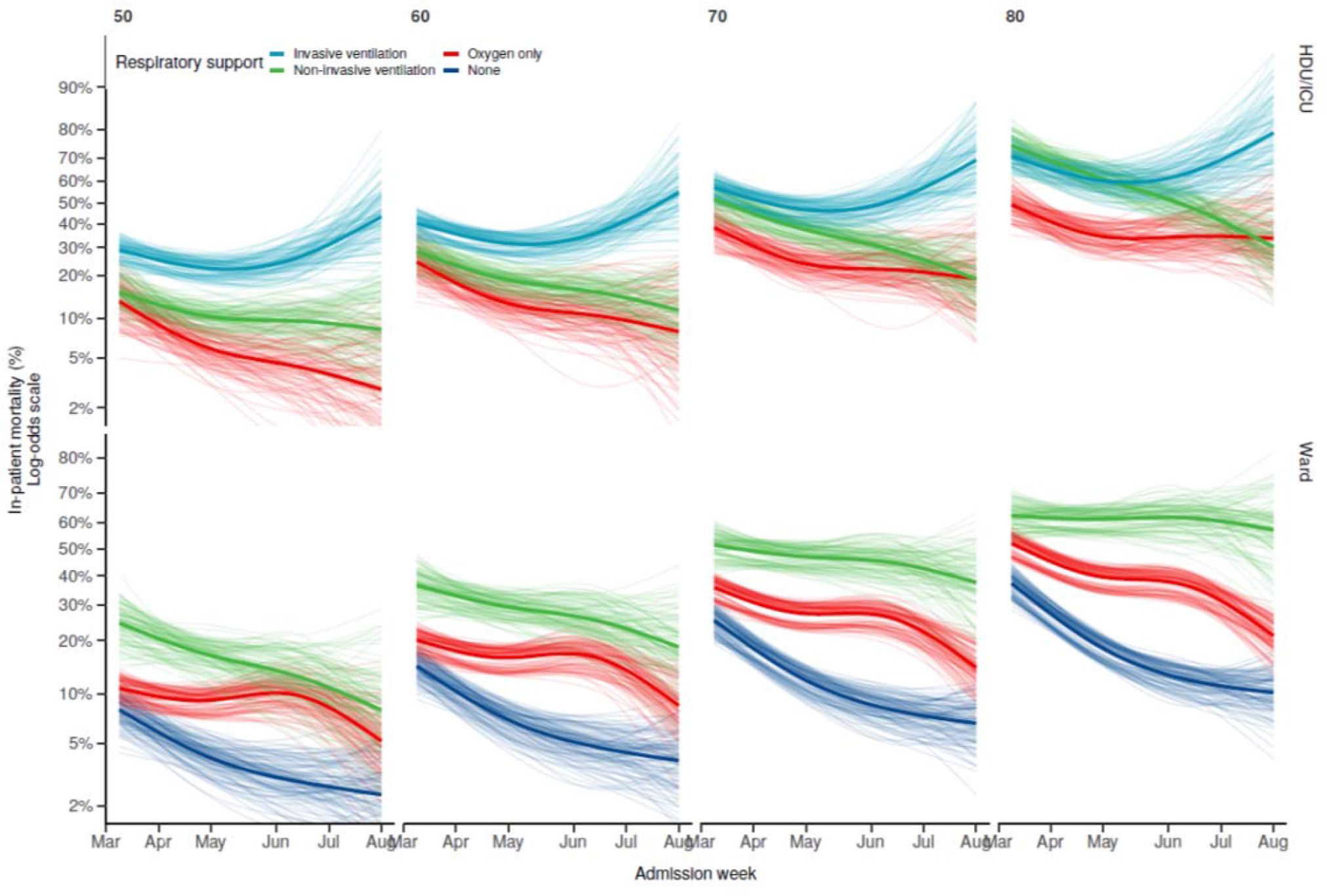
Mortality in adults admitted to hospital with COVID-19 stratified by respiratory support and age. Bayesian generalised additive model, adjusted for age, sex, comorbidity, GCS, respiratory rate, SpO2, serum urea and CRP, with 3-way interaction between age, week of admission, and level of respiratory support. Missing data imputed with 10 datasets. Plot lines represent samples from posterior distribution.

**Figure 6:**
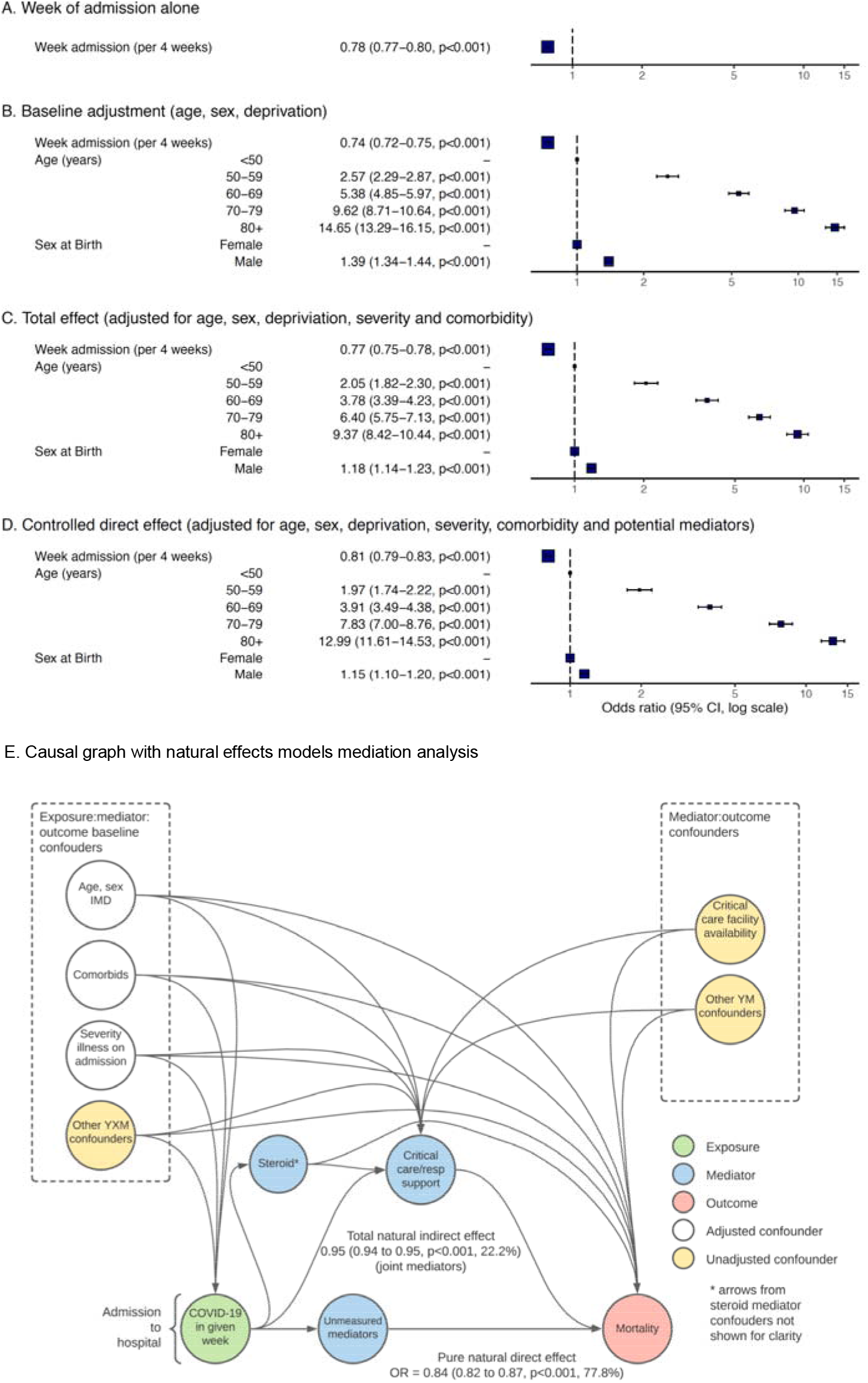
Odds ratio for hospital mortality for week of admission. A: unadjusted week of admission, B: adjusted for age, sex and deprivation, C: in addition adjusted for comorbidity count, deprivation, illness severity (respiratory rate, oxygen saturations, GCS, serum Urea, CRP), D: in addition adjusted for mediators (icu/ward:respiratory support * steroid). E: Total natural indirect effect OR 0.95 (0.94 to 0.96, p<0.001, 22.2%) (joint mediators). Pure natural direct effect OR 0.84 (0.82 to 0.87, p<0.001, 77.8%).

There were significant reductions in unadjusted mortality between TP1 and TP3 in patients receiving no respiratory support (TP1 14.4% vs TP3 6.0%), oxygen only (Ward: 35.1% vs 21.3%; Critical Care 22.6% vs 12.8%), and NIV in ICU (33.3% vs 24.8%) across all age groups (Figure 4, Table E10). However, mortality remained persistently high for patients receiving IMV (41.1% vs 41.8%) and NIV on the ward (48.0% vs 44.3%). These differential changes in mortality persisted after adjustment for demographic and severity of illness variables (Figure 5).

## Discussion

Overall hospital mortality within 28 days after admission substantially decreased throughout the course of the first wave. At the peak of admissions in late March and early April, illness severity at hospital presentation was greatest, and patients presented later from their onset of symptoms. There was a reduction in the level of respiratory support received: use of invasive ventilation reduced over time, and the proportion of non-invasive ventilation increased. By late June/July, nearly half of patients admitted required no supplementary oxygen. The reduction in hospital mortality was seen across all demographic groups, and was not fully accounted for by the fall in case mix or illness severity. One fifth of the reduction in mortality could be accounted for by changes in treatment including respiratory care and steroids.

ISARIC4C has recruited patients across the UK, accounting for approximately two-thirds of patients admitted to hospital in the UK with COVID-19 in the first wave. Data were collected from the front door to discharge for patients managed both on the ward and within critical care enabling us to review admissions and mortality for the whole hospital. We did not record treatment escalation plans, but we were able to examine changing case-mixes in the ward and critical care. Due to the nature of the pandemic, there were more missing data than would normally be expected in a prospective cohort study, but this was handled using appropriate methods. We were unable to comment on community factors leading up to admission, and indeed those who were not admitted to hospital. This was an observational study, as such we were unable to assign causality and unmeasured confounding may remain. The issue of conditioning on a collider for critical care cohort is well established in critical care epidemiology literature and its impact on associations is understood. This comparison of ward vs critical care cohorts is absent from most other literature.

We have demonstrated a reduction in hospital mortality that cannot be fully explained by baseline demographics or measured presenting severity of illness markers. This is consistent with experiences in New York hospitals^20^ where mortality also significantly and progressively fell over the course of the study period. Critical care mortality in the UK has also reduced.^21,22^ The majority of patients admitted to hospital throughout the first wave were elderly, comorbid and of White ethnicity, and these groups had the highest mortality. Case mix changed over the pandemic, with a rise in the proportion of younger and female patients who, both in our study and others, have lower mortality rates.^23,24^ However the falls in mortality were seen in all ages, ethnic groups, and in both sexes. Shielding of vulnerable groups was formally introduced on 23 March^25^ and earlier patients may have been more vulnerable but not identifiable in our dataset. Severity of illness at presentation to hospital fell and increasingly patients required no respiratory support at all. SARS-CoV-2 infection is transmitted predominantly by respiratory droplets, so social distancing, UK lockdown (March 23 2020)^26^, and widespread adoption of masks may have reduced viral load (infectious dose) at point of transmission^27^, in turn reducing severity of illness in infected patients.^28^ Patients presented earlier in their disease course in later months and length of stay for non-survivors in critical care increased consistent with patients presenting earlier in their illness, and less in-extremis. Our data do not include community factors, however it may be that changes in health seeking behaviour enabled patients to attend hospitals more easily.

Hospital admissions in the UK peaked at approximately 3,000 patients admitted to hospital each day in early April, with Intensive Care Unit (ICU) caseload peaking shortly after,^1,2^ gradually falling to a plateau of approximately 100 patients per day by July.^1^ The UK has a small health care workforce, with relatively few hospital and critical care beds in comparison to other high income countries.^29,30^ Coming into the pandemic, the UK had high levels of bed occupancy and very little spare capacity.^29^ However, during the rapid response to COVID-19 there was a substantial increase in capacity and national reported occupancy for critical care never exceeded 60%, although local peaks were much higher.^20^ Even during more normal times, critical care capacity strain is associated with increased mortality.^31^ The rapid expansion of critical care beds required redeployment of non-critical care staff, and in some UK regions, increased ratios of nursing staff to patients which may have impacted on early patient outcomes.^32^

During the peak of admissions, and during peak illness severity, a higher proportion of patients were admitted to critical care. Patients in critical care at this time were considerably younger than ward patients, even so, mortality was much higher than for other SARIs such as viral pneumonia.^5^ During the peak admission period, the proportion of patients aged >80 years and those with multimorbidity admitted to critical care was lower than subsequently, however, this reflected the demographic pattern also seen in patients admitted to the ward at this time.

A fifth of the fall in mortality can be explained by changes in respiratory support and steroids along with associated changes in clinical decision-making relating to the former intervention. The proportion of patients receiving invasive ventilation reduced over time, however mortality remained persistently high after adjusting for demographics and illness severity. It would be unwise to interpret this association as causal as use of IMV is reserved for those with the most severe illness and, overall, mortality reduced for patients admitted to critical care. There are several potential explanations for this finding. Firstly, the change in case mix may not have been adequately captured by multi-morbidity and age: a higher proportion of more elderly and comorbid patients were ventilated later in the pandemic, potentially when there was more critical care capacity. Secondly, practice within critical care changed, with increasing use of NIV over time, and only those presenting in extremis or failing a trial of NIV received IMV. This may be partly due to the changing case mix, but also due to increasing clinician familiarity with the use of NIV, and an improving ability to identify which patients might benefit. Potentially, early patients who received IMV would later have received NIV and have survived regardless of the mode of ventilation. The later patients receiving IMV were therefore a more severely ill population who had failed to respond to treatments, and would die if not offered IMV. This is supported by the significant increase in the effect of respiratory care over time: allocation to respiratory support was linked to better prediction of outcome by clinicians over time. Ongoing trials comparing the use of NIV and IMV in critically ill patients with COVID-19 will be able to overcome this selection bias and confounding by indication to answer whether patient selection or NIV itself is improving outcomes in critically ill patients with COVID-19.^33^

Patients receiving NIV on the ward had higher mortality rates than patients on oxygen and NIV managed in critical care. Along with the higher rates of comorbidity, in particular dementia and chronic pulmonary and cardiac disease, this may reflect that the benefit of these treatments is limited in this group of patients, as well as a potential ceiling of treatment for patients receiving NIV on the ward. Mortality was extremely high in elderly patients who received invasive ventilation. The benefit of ICU admission for elderly frail patients remains uncertain as this is a population with high rates of mortality and long-term functional impairment in survivors.^34^ Critical care interventions may not be associated with improved outcomes in this group: in a previous study, protocolised ICU referral in the elderly led to significantly higher ICU admission rates, but without a significant effect on mortality, functional status or health-related quality of life.^35^ It is essential that meaningful discussions about the treatment options available as well as their risks and benefits are discussed with these patients and their families.^36^ Such discussions should also emphasise that much of the potential benefit of care can be derived without the need for ICU level care.

Practice outside critical care has also changed, with increasing clinical familiarity with COVID-19. Clinicians may have become more alert to deterioration, which can occur rapidly in COVID-19 and may not have been accurately captured by our data collection. Corticosteroid treatment^7,8^ substantially benefits subgroups of the hospital population, and trials of other treatments including anticoagulation, anti-inflammatory and anti-viral agents, convalescent plasma and non-invasive ventilation are ongoing.^7,8^ This highlights the critical importance of suitably-powered randomised controlled trials for drug evaluation even in outbreak situations.

## Conclusion

In-hospital mortality rates for patients with COVID-19 fell in the UK over the course of the first wave. This fall persisted after adjusting for illness severity and changes in patient case-mix. Patients were most severely unwell at hospital presentation at the start of the pandemic and presented later in their disease course. A significant proportion of the fall may be explained by changes in management including respiratory support and steroid treatment. Hospital practice has changed, in particular the use of NIV increased dramatically, and many patients have been included in drug and other treatment trials, which may help to explain the fall in mortality and inform future waves. Hospital mortality remained high for patients receiving invasive ventilation and ward non-invasive ventilation, and these populations should be a priority in ongoing research.

## Declarations

ABD, RHM and EMH verify the underlying data.

### Ethics

Ethical approval for data collection and analysis by ISARIC4C was given by the South Central-Oxford C Research Ethics Committee in England (reference 13/SC/0149), and by the Scotland A Research Ethics Committee (reference 20/SS/0028). The ISARIC WHO CCP-UK study was registered at https://www.isrctn.com/ISRCTN66726260 and designated an Urgent Public Health Research Study by NIHR.

## Data Availability

This work uses data provided by patients and collected by the NHS as part of their care and support #DataSavesLives. The CO-CIN data was collated by ISARIC4C Investigators. ISARIC4C welcomes applications for data and material access through our Independent Data and Material Access Committee (https://isaric4c.net).

## Acknowledgments

This work uses data provided by patients and collected by the NHS as part of their care and support #DataSavesLives. We are extremely grateful to the 2,648 frontline NHS clinical and research staff and volunteer medical students, who collected this data in challenging circumstances; and the generosity of the participants and their families for their individual contributions in these difficult times. We also acknowledge the support of Jeremy J Farrar and Nahoko Shindo.

## Author contributions

Conceptualisation: JK Baillie, J Dunning, JS Nguyen-Van-Tam, PJM Openshaw, MG Semple. Formal analysis: AB Docherty, RH Mulholland, NI Lone, EM Harrison, MG Semple, C Cheyne, D De Angelis, K Diaz-Ordaz, S Funk, M Garcia-Finana, D Hughes, R Keogh, PD Kirwan. Writing original draft: AB Docherty, RH Mulholland, NI Lone, EM Harrison, MG Semple. Writing reviewing and editing: JK Baillie, AB Docherty, TM Drake, J Dunning, EM Harrison, A Ho, JS Nguyen-Van-Tam, Clark D Russell, BDM Tom, L Turtle, PJM Openshaw, R Pius, MG Semple. Project administration: C Donoghue HE Hardwick, A Ho, M Girvan, J Harrison, G Leeming, R Spencer. Funding acquisition: JK Baillie, PJM Openshaw, MG Semple.

## References

1. GOV.UK. Coronavirus (COVID-19) in the UK. Available from: https://coronavirus.data.gov.uk/ [Accessed 18 November 2020].

2. Heneghan C, Oke J. COVID-19: Admissions to Hospital – Update. The Centre for Evidence-Based Medicine. 2020. Available from: https://www.cebm.net/covid-19/covid-19-uk-hospital-admissions/. [Accessed 18 November 2020].

3. Mahon J, Oke J, Heneghan C. Declining death rate from COVID-19 in hospitals in England. The Centre for Evidence-Based Medicine. 2020. [Accessed 18 November 2020].

4. Zeffman H. Doctors report big drop in hospital coronavirus deaths. The Times. 2020.

5. ICNARC (Intensive Care National Audit and Research Centre). Reports. Available from: https://www.icnarc.org/Our-Audit/Audits/Cmp/Reports [Accessed 24 November 2020]

6. SICSAG (Scottish Intensive Care Society). Publications. Available from: https://www.sicsag.scot.nhs.uk/publications/main.htm [Accessed 24 November 2020]

7. RECOVERY Collaborative Group. Dexamethasone in Hospitalized Patients with Covid-19 — Preliminary Report. The New England Journal of Medicine. Available from: doi: 10.1056/NEJMoa2021436 [Accessed 24 November 2020].

8. REMAP-CAP Investigators. Effect of Hydrocortisone on Mortality and Organ Support in Patients With Severe COVID-19. JAMA. 2020. 324(13):1317–1329. Available from: doi:10.1001/jama.2020.17022

9. Dunning J.W, Merson L., Rohde G.G.U et al.. Open source clinical science for emerging infections. The Lancet Infectious Diseases. 2014. 14 (1): 8–9. Available from: https://doi.org/10.1016/S1473-3099(13)70327-X

10. ISARIC-4C Investigators. Available from: https://isaric4c.net/. [Accessed 18 November 2020].

11. Docherty A.B, Harrison E.M, Hardwick H.E et al. Features of 201133 UK patients in hospital with covid-19 using the ISARIC WHO Clinical Characterisation Protocol: prospective observational cohort study. BMJ. 2020. 369:m1985. Available from: doi:10.1136/bmj.m1985.

12. Knight S.R, Ho A, Buchan I et al. Risk stratification of patients admitted to hospital with covid-19 using the ISARIC WHO Clinical Characterisation Protocol: development and validation of the 4C Mortality Score. BMJ. 2020. 370:m3339. Available from: doi:10.1136/bmj.m1985.

13. GOV.UK. COVID-19: Government announces moving out contain phase into delay. 2020. Available from: https://www.gov.uk/government/news/covid-19-government-announces-moving-out-of-contain-phase-and-into-delay

14. NHS Health Research Authority. Extension of COVID-19 COPI Notice. 2020. Available from: https://www.hra.nhs.uk/about-us/news-updates/extension-covid-19-copi-notice/

15. Public Benefit and Privacy Panel for Health and Social Care. Available from: https://www.informationgovernance.scot.nhs.uk/pbpphsc/

16. Harrison E.M, Annemarie A.B, Barr B, et al. Ethnicity and Outcomes from COVID-19: The ISARIC CCP-UK Prospective Observational Cohort Study of Hospitalised Patients. SSRN. 2020. Available from: doi: 10.2139/ssrn.3618215.

17. The WHO Rapid Evidence Appraisal for COVID-19 Therapies (REACT) Working Group. Association Between Administration of Systemic Corticosteroids and Mortality Among Critically Ill Patients With COVID-19. A Meta-analysis. JAMA. 2020. 324(13):1330–1341. Available from: doi:10.1001/jama.2020.17023.

18. Griffin S. Covid-19: England comes into line with rest of UK on recording deaths. BMJ. 2020. 370:m3220. Available from: doi:10.1136/bmj.m3220.

19. Vansteelandt A, Bakaert M, Lange T. Imputation Strategies for the Estimation of Natural Direct and Indirect Effects. Epidemiologic Methods. 2012. 1 (1). Available from: doi: 10.1515/2161-962X.1014

20. Horwitz L.I, Jones S.A, Cerfolio R.J, et al. Trends in COVID-19 Risk-Adjusted Mortality Rates. Journal of Hospital Medicine. 2020. Available from: doi:10.12788/jhm.3552.

21. Doidge J.C, Mouncey P.R, Thomas K, et al. Trends in Intensive Care for Patients with COVID-19 in England, Wales and Northern Ireland. Preprints. 2020. 2020080267. Available from: doi:10.20944/preprints202008.0267.v1.

22. Dennis J.M, McGovern A.P, Vollmer S.J, Mateen B.A. Improving Survival of Critical Care Patients With Coronavirus Disease 2019 in England. Critical Care Medicine. 2020. Available from: doi: 10.1097/CCM.0000000000004747.

23. WHO. Coronavirus Disease (COVID-19). October 2020. Available from: https://apps.who.int/iris/bitstream/handle/10665/336034/nCoV-weekly-sitrep11Oct20-eng.pdf

24. Bonanad C, Garcia-Blas S, Tarazona-Santabalbina F et al. The Effect of Age on Mortality in Patients With COVID-19: A Meta-Analysis With 611,583 Subjects. Journal of the American Medical Directors Association. 2020. 21, 7, 915–918. Available from: doi:10.1016/j.jamda.2020.05.045

25. Department of Health and Social Care. Shielding letter to NHS. 2020. Available from: https://www.england.nhs.uk/coronavirus/wp-content/uploads/sites/52/2020/06/C0624-shielding-letter-to-nhs.pdf

26. GOV.UK. Prime Minister’s statement on coronavirus (COVID-19): 23 March 2020. Available from: https://www.gov.uk/government/speeches/pm-address-to-the-nation-on-coronavirus-23-march-2020

27. Goyal A, Reeves D.B, Cardozo-Ojeda E.F, Mayer B.T, Schliffer J.T. Slight reduction in SARS-CoV-2 exposure viral load due to masking results in a significant reduction in transmission with widespread implementation. medRxiv. 2020. Available from: doi: 10.1101/2020.09.13.20193508.

28. Pujadas E, Chaudhry F, McBride R, et al. SARS-CoV-2 viral load predicts COVID-19 mortality. The Lancet Respiratory Medicine. 2020. Available from: doi: doi.org/10.1016/S2213-2600(20)30354-4.

29. Thomas C. Resilient Health and Care. Learning the lessons of COVID-19 in the English NHS. Institute for Public Policy Research. 2020. Available from: https://www.ippr.org/files/2020-07/resilient-health-and-care-july20.pdf

30. OECD. Intensive care beds capacity. 2020. Available from: https://www.oecd.org/coronavirus/en/data-insights/intensive-care-beds-capacity

31. Harris S, Singer M, Sanderson C, Grieve R, Harrison D, Rowan K. Impact on mortality of prompt admission to critical care for deteriorating ward patients: an instrumental variable analysis using critical care bed strain. Intensive Care Medicine. 2018. 11. 606–615. Available from: doi: https://doi.org/10.1007/s00134-018-5148-2

32. Sprivulis P.C, Da Silva J.A, Jacobs I.G, Jelinek G.A, Frazer A.R.L. The association between hospital overcrowding and mortality among patients admitted via Western Australian emergency departments. MJA. 2006. 184, 5, 208–212. Available from: doi: 10.5694/j.1326-5377.2006.tb00203.x

33. RECOVERY Collaborative Group. RECOVERY-RS Respiratory Support : Respiratory Strategies in COVID-19; CPAP, High-flow, and standard care. Warwick Clinical Trials Unit. 2020. Available from: https://warwick.ac.uk/fac/sci/med/research/ctu/trials/recovery-rs

34. Guillion A, Hermetet C, Barker K.A et al. Long-term survival of elderly patients after intensive care unit admission for acute respiratory infection: a population-based, propensity score-matched cohort study. Critical Care. 2020. 24, 384. Available from: doi: 10.1186/s13054-020-03100-4.

35. Guidet B, Leblanc G, Simon T et al. on behalf of the ICE-CUB 2 study network. Effect of systematic intensive care unit triage on long-term mortality among critically ill elderly patients in France: a randomized clinical trial. JAMA. 2017. 318. 1450–1459. Available from: doi: 10.1001/jama.2017.13889

36. Realistic Medicine. About. Available from: https://www.realisticmedicine.scot/about/.

