## Supplementary material for "Changes in UK hospital mortality in the first wave of COVID-19: the ISARIC WHO Clinical Characterisation Protocol prospective multicentre observational cohort study": Online supplement

### Supplementary info

#### Definitions

| Term | Definition |
|---|---|
| Acute hospitals | Provides general short-term healthcare treatment such as treatment for severe injury, period of illness, urgent medical condition, or to recover from surgery. In the NHS, it often includes services such as accident and emergency (A&E) departments, inpatient and outpatient medicine and surgery |
| Non-invasive ventilation (NIV) | bilevel and continuous positive airway pressure |
| Invasive mechanical ventilation (IMV) | Positive pressure ventilation delivered via a tube inserted into the trachea (through the mouth, nose, or skin in the neck) |
| Level of care | "Ward": Level 0/1<br>- Ward based care where the patient does not require organ support (for example, they may need an IV, or oxygen by face mask)<br>Critical Care: Level 2 or 3<br>- Level 2: High dependency unit (HDU). Patients needing single organ support (excluding mechanical ventilation) such as renal haemofiltration or ionotropes and invasive BP monitoring. They are staffed with one nurse to two patients<br>- Level 3: Intensive care (ICU). Patients requiring two or more organ support (or needing mechanical ventilation alone). Staffed with one nurse per patient and usually with a doctor present in the unit 24 hours per day. |

#### Exclusions

For this analysis, we included only acute general hospitals. Community hospitals providing long-term treatment and residential mental health hospitals were excluded since patient populations were very different and were therefore not comparable. We excluded 'nosocomial' infection as patients with onset of COVID-19 symptoms more than 5 days after they were admitted to hospital for a separate condition.[1] We excluded patients where data entries for outcome date and age were clearly erroneous. Onset of symptoms to admission ≥21 days were assumed to be errors.

Site training emphasised that only patients who tested positive (using reverse transcriptase polymerase chain reaction, RT-PCR) for COVID-19 were eligible for enrolment.

#### Variables

Comorbidities: asthma, diabetes, chronic cardiac disease (excluding hypertension), chronic haematologic disease, chronic kidney disease, chronic neurological disorder, chronic pulmonary disease (excluding asthma), dementia, HIV/AIDS, malignancy, malnutrition, mild to severe liver disease, clinician-assigned obesity and rheumatologic disorder.

---

[1] Rickman H.M, Rampling T, Shaw K et al. Nosocomial Transmission of Coronavirus Disease 2019: A Retrospective Study of 66 Hospital-acquired Cases in a London Teaching Hospital. *Clinical Infectious Diseases.* 2020. ciaa816. Available from: doi:1093/cid/ciaa816.

#### Statistical methods

Secondary outcome: A Bayesian approach was taken to facilitate the use of imputed datasets with GAMs, and to allow the easy determination of probabilities of interest. Continuous and binary data were centred and normalised. Weakly informative priors were used for regression coefficients (Student's t-distribution, df = 7, location = 0, scale = 2.5) and for the standard deviation of smooth terms (exponential, rate = 1). Each imputed data set (n = 10) was fitted separately (chains = 4, total iterations = 5000, warm-up = 1000, random initiation values from -2 to 2) and results pooled across models. Posterior distributions were sampled from pooled data. Convergence was ensured by inspecting chain trace, density, and autoregression plots, and determining the Gelman/Rubin potential scale reduction factor (R-hat).

Models incorporated variables previously shown to be associated with in-hospital mortality: age, sex, comorbidity count, and severity of illness (RR, SpO2, GCS, urea, CRP)). Based on our previous work, a three-way interaction was included between admission week, age, and maximum level of respiratory support. Multivariable GLMs were used to fit in-hospital mortality to week of admission (straight lines on log odds scale). Multivariable GAMs using restricted cubic splines were then used to allow the response to vary for all continuous variables. Admission week and age smooth terms were allowed to vary by maximum respiratory support and a tensor product smooth term used to model the admission week and age interaction.

Statistical disclosure control (SDC) measures were adopted in the summary statistics tables to reduce the risk of patient confidentiality. If cell was <5 then it was eitherremoved (and replaced with NA) or aggregated with another level to hide small numbers. Note NA could be suppressed or 0. If small cell was an 'Unknown' category these were left as we are assuming they are random by chance.

### Tables

Table E1: Individual comorbidities of patients presenting in the first wave, divided by 3 equal time points.

| **Comorbidity** | **1 - Weeks 11 to 17** 9 Mar to 26 Apr 2020 | **2 - Weeks 18 to 24** 27 Apr to 14 Jun 2020 | **3 - Weeks 25 to 31** 15 Jun to 2 Aug 2020 |
|---|---|---|---|
| **AIDS/HIV** | 178 (0.4%) | 41 (0.3%) | 13 (0.5%) |
| **Asthma** | 6373 (13.4%) | 1612 (11.7%) | 336 (12.1%) |
| **Chronic cardiac disease** | 13837 (29.2%) | 4745 (34.5%) | 883 (31.8%) |
| **Chronic hematologic disease** | 1833 (3.9%) | 633 (4.6%) | 118 (4.3%) |
| **Chronic kidney disease** | 60 (15.7%) | 2551 (18.6%) | 483 (17.4%) |
| **Chronic neurological disorder** | 5338 (11.2%) | 1829 (13.3%) | 304 (11%) |
| **Chronic pulmonary disease** | 7773 (16.4%) | 2482 (18.1%) | 482 (17.4%) |
| **Dementia** | 6992 (14.7%) | 2666 (19.4%) | 343 (12.4%) |
| **Diabetes Type** | | | |
| Type 1 | 720 (1.5%) | 393 (2.9%) | 66 (2.4%) |
| Type 2 | 6166 (13%) | 3275 (23.8%) | 659 (23.7%) |
| **Hypertension*** | 11061 (23.3%) | 6133 (44.6%) | 1185 (42.7%) |
| **Malignant neoplasm** | 4167 (8.8%) | 1495 (10.9%) | 315 (11.4%) |
| **Malnutrition** | 988 (2.1%) | 375 (2.7%) | 68 (2.5%) |
| **Mild to severe liver disease** | 1342 (2.8%) | 527 (3.8%) | 104 (3.7%) |
| **Obesity** | 4923 (10.4%) | 1342 (9.8%) | 294 (10.6%) |
| **Rheumatologic disorder** | 4804 (10.1%) | 1674 (12.2%) | 308 (11.1%) |

* Hypertension added May 2020

Table E2: Baseline characteristics of ICU patients presenting in the first wave, divided by 3 equal time points (N=9,340). Disclosure threshold of 5 was used.

| Type | Characteristic | 1 - Weeks 11 to 17<br>9 Mar to 26 Apr 2020 | 2 - Weeks 18 to 24<br>27 Apr to 14 Jun 2020 | 3 - Weeks 25 to 31<br>15 Jun to 2 Aug 2020 |
|---|---|---|---|---|
| | **Total** | 7732 | 1275 | 333 |
| Patient characteristics | **Country** | | | |
| | England | 6973 (90.2%) | 1153 (90.4%) | 312 (93.7%) |
| | Scotland | 460 (5.9%) | 53 (4.2%) | NA |
| | Wales | 299 (3.9%) | 69 (5.4%) | 21 (6.3%) |
| | **Age (grouped)** | | | |
| | <50 | 1564 (20.2%) | 266 (20.9%) | 79 (23.7%) |
| | 50-69 | 4194 (54.2%) | 637 (50%) | 146 (43.8%) |
| | 70-79 | 1499 (19.4%) | 244 (19.1%) | 69 (20.7%) |
| | 80+ | 475 (6.1%) | 128 (10%) | 39 (11.7%) |
| | **Age (continuous)** | | | |
| | Median (IQR) | 61 (IQR=17.9) | 62 (IQR=20.4) | 64 (IQR=22.4) |
| | Mean (SD) | 60.6 (SD=13.4) | 61.3 (SD=15) | 61.7 (SD=16.1) |
| | **Sex** | | | |
| | Female | 2314 (29.9%) | 465 (36.5%) | 128 (38.4%) |
| | Male | 5418 (70.1%) | 810 (63.5%) | 205 (61.6%) |
| | **Ethnic group** | | | |
| | White | 4754 (61.5%) | 869 (68.2%) | 209 (62.8%) |
| | South Asian | 562 (7.3%) | 87 (6.8%) | 54 (16.2%) |
| | East Asian | 124 (1.6%) | 10 (0.8%) | NA |
| | Black | 468 (6.1%) | 57 (4.5%) | 8 (2.4%) |
| | Other Ethnic Minority | 831 (10.7%) | 112 (8.8%) | 40 (12%) |
| | Unknown | 993 (12.8%) | 140 (11%) | 22 (6.6%) |
| | **Number of comorbidities** | | | |
| | 0 | 2655 (34.3%) | 259 (20.3%) | 57 (17.1%) |
| | 1 | 2238 (28.9%) | 318 (24.9%) | 80 (24%) |
| | 2+ | 2839 (36.7%) | 698 (54.7%) | 196 (58.9%) |
| | **AIDS/HIV** | 54 (0.7%) | 9 (0.7%) | NA |
| | **Asthma** | 1242 (16.1%) | 187 (14.7%) | 54 (16.2%) |
| | **Chronic cardiac disease** | 1147 (14.8%) | 250 (19.6%) | 76 (22.8%) |
| | **Chronic hematologic disease** | 222 (2.9%) | 34 (2.7%) | 10 (3%) |
| | **Chronic kidney disease** | 72 (7.4%) | 129 (10.1%) | 35 (10.5%) |
| | **Chronic neurological disorder** | 375 (4.8%) | 87 (6.8%) | 25 (7.5%) |
| | **Chronic pulmonary disease** | 693 (9%) | 172 (13.5%) | 61 (18.3%) |
| | **Dementia** | 141 (1.8%) | 45 (3.5%) | 5 (1.5%) |
| | **Diabetes Type** | | | |
| | Type 1 | 109 (1.4%) | 49 (3.8%) | 11 (3.3%) |
| | Type 2 | 954 (12.3%) | 343 (26.9%) | 78 (23.4%) |
| | **Hypertension** | 1512 (19.6%) | 522 (40.9%) | 138 (41.4%) |
| | **Malignant neoplasm** | 369 (4.8%) | 101 (7.9%) | 20 (6%) |
| | **Malnutrition** | 71 (0.9%) | 19 (1.5%) | NA |
| | **Mild to severe liver disease** | 172 (2.2%) | 45 (3.5%) | 13 (3.9%) |
| | **Obesity** | 1524 (19.7%) | 282 (22.1%) | 71 (21.3%) |
| | **Rheumatologic disorder** | 531 (6.9%) | 102 (8%) | 21 (6.3%) |
| | **Health worker** | 682 (8.8%) | 177 (13.9%) | 15 (4.5%) |
| Severity of illness | **Asymptomatic** | 184 (2.4%) | 91 (7.1%) | 39 (11.7%) |
| | **Symptom onset (days)*** | | | |
| | Median (IQR) | 7 (IQR=7) | 5 (IQR=7) | 4 (IQR=6) |
| | Mean (SD) | 7 (SD=4.9) | 5.5 (SD=5.1) | 4.9 (SD=4.6) |
| | **Length of stay (days)** | | | |
| | Median (IQR) | 14 (IQR=18) | 14 (IQR=17) | 11 (IQR=13) |
| | Mean (SD) | 19.7 (SD=17.5) | 17.9 (SD=14.9) | 13.8 (SD=10.1) |
| | **ISARIC 4C Score** | | | |
| | Low (0-3) | 215 (2.8%) | 62 (4.9%) | 17 (5.1%) |
| | Intermediate (4-8) | 1797 (23.2%) | 324 (25.4%) | 90 (27%) |
| | High (9-14) | 2677 (34.6%) | 483 (37.9%) | 122 (36.6%) |
| | Very high (15+) | 492 (6.4%) | 89 (7%) | 20 (6%) |
| | Unknown | 2551 (33%) | 317 (24.9%) | 84 (25.2%) |

| | | | | |
|---|---|---|---|---|
| | **Respiratory rate (breaths/min)** | | | |
| | <20 | 1317 (17%) | 285 (22.4%) | 68 (20.4%) |
| | 20-30 | 3737 (48.3%) | 640 (50.2%) | 170 (51.1%) |
| | >=30 | 2379 (30.8%) | 315 (24.7%) | 89 (26.7%) |
| | Unknown | 299 (3.9%) | 35 (2.7%) | 6 (1.8%) |
| | **Peripheral oxygen saturation on room air (%)** | | | |
| | >=92 | 4552 (58.9%) | 841 (66%) | 221 (66.4%) |
| | <92 | 2918 (37.7%) | 408 (32%) | 105 (31.5%) |
| | Unknown | 262 (3.4%) | 26 (2%) | 7 (2.1%) |
| | **Glasgow coma score** | | | |
| | 15 | 5212 (67.4%) | 973 (76.3%) | 256 (76.9%) |
| | <15 | 1484 (19.2%) | 211 (16.5%) | 45 (13.5%) |
| | Unknown | 1036 (13.4%) | 91 (7.1%) | 32 (9.6%) |
| | **Urea (mmol/L)** | | | |
| | <7 | 3316 (42.9%) | 603 (47.3%) | 181 (54.4%) |
| | 7-14 | 2110 (27.3%) | 346 (27.1%) | 82 (24.6%) |
| | >14 | 831 (10.7%) | 162 (12.7%) | 30 (9%) |
| | Unknown | 1475 (19.1%) | 164 (12.9%) | 40 (12%) |
| | **C-reactive protein (mg/dL)** | | | |
| | <50 | 888 (11.5%) | 279 (21.9%) | 90 (27%) |
| | 50-99 | 1269 (16.4%) | 245 (19.2%) | 67 (20.1%) |
| | >=100 | 4438 (57.4%) | 609 (47.8%) | 132 (39.6%) |
| | Unknown | 1137 (14.7%) | 142 (11.1%) | 44 (13.2%) |
| **Respiratory support and treatments** | **Respiratory support** | | | |
| | Oxygen only | 1006 (13%) | 260 (20.4%) | 78 (23.4%) |
| | Non-invasive | 1768 (22.9%) | 461 (36.2%) | 157 (47.1%) |
| | Invasive | 4958 (64.1%) | 554 (43.5%) | 98 (29.4%) |
| | **Proning** | 3110 (40.2%) | 402 (31.5%) | 61 (18.3%) |
| | **Extra corporeal memberance oxygenation (ECMO)** | 267 (3.5%) | 41 (3.2%) | 6 (1.8%) |
| | **Steroids** | | | |
| | Yes | 1667 (21.6%) | 378 (29.6%) | 217 (65.2%) |
| | No | 1779 (23%) | 433 (34%) | 89 (26.7%) |
| | Unknown | 4286 (55.4%) | 464 (36.4%) | 27 (8.1%) |

* Symptom onset summary statistics based on patients with symptoms up to 3 weeks before admission only

Table E3: Baseline characteristics of ward patients presenting in the first wave, divided by 3 equal time points (N=54,632). Disclosure threshold of 5 was used.

| Type | Characteristic | 1 - Weeks 11 to 17<br>9 Mar to 26 Apr 2020 | 2 - Weeks 18 to 24<br>27 Apr to 14 Jun 2020 | 3 - Weeks 25 to 31<br>15 Jun to 2 Aug 2020 |
|---|---|---|---|---|
| | **Total** | 39721 | 12469 | 2442 |
| Patient characteristics | **Country** | | | |
| | England | 36353 (91.5%) | 11426 (91.6%) | 2277 (93.2%) |
| | Scotland | 1680 (4.2%) | 481 (3.9%) | 33 (1.4%) |
| | Wales | 1688 (4.2%) | 562 (4.5%) | 132 (5.4%) |
| | **Age (grouped)** | | | |
| | <50 | 4697 (11.8%) | 1397 (11.2%) | 407 (16.7%) |
| | 50-69 | 10090 (25.4%) | 2624 (21%) | 588 (24.1%) |
| | 70-79 | 9176 (23.1%) | 2771 (22.2%) | 521 (21.3%) |
| | 80+ | 15758 (39.7%) | 5677 (45.5%) | 926 (37.9%) |
| | **Age (continuous)** | | | |
| | Median (IQR) | 76 (IQR=23.5) | 78.1 (IQR=22) | 74.87 (IQR=25.7) |
| | Mean (SD) | 72 (SD=16.7) | 73.7 (SD=17.1) | 70 (SD=18.7) |
| | **Sex** | | | |
| | Female | 17523 (44.1%) | 6194 (49.7%) | 1225 (50.2%) |
| | Male | 22198 (55.9%) | 6275 (50.3%) | 1217 (49.8%) |
| | **Ethnic group** | | | |
| | White | 29239 (73.6%) | 9963 (79.9%) | 1713 (70.1%) |
| | South Asian | 1717 (4.3%) | 443 (3.6%) | 242 (9.9%) |
| | East Asian | 274 (0.7%) | 31 (0.2%) | NA |
| | Black | 1547 (3.9%) | 179 (1.4%) | 33 (1.4%) |
| | Other Ethnic Minority | 2509 (6.3%) | 569 (4.6%) | 201 (8.2%) |
| | Unknown | 4435 (11.2%) | 1284 (10.3%) | 253 (10.4%) |
| | **Number of comorbidities** | | | |
| | 0 | 8134 (20.5%) | 1755 (14.1%) | 450 (18.4%) |
| | 1 | 9151 (23%) | 2145 (17.2%) | 466 (19.1%) |
| | 2+ | 22436 (56.5%) | 8569 (68.7%) | 1526 (62.5%) |
| | **AIDS/HIV** | 124 (0.3%) | 32 (0.3%) | NA |
| | **Asthma** | 5131 (12.9%) | 1425 (11.4%) | 282 (11.5%) |
| | **Chronic cardiac disease** | 12690 (31.9%) | 4495 (36%) | 807 (33%) |
| | **Chronic hematologic disease** | 1611 (4.1%) | 599 (4.8%) | 108 (4.4%) |
| | **Chronic kidney disease** | 6888 (17.3%) | 2422 (19.4%) | 448 (18.3%) |
| | **Chronic neurological disorder** | 4963 (12.5%) | 1742 (14%) | 279 (11.4%) |
| | **Chronic pulmonary disease** | 7080 (17.8%) | 2310 (18.5%) | 421 (17.2%) |
| | **Dementia** | 6851 (17.2%) | 2621 (21%) | 338 (13.8%) |
| | **Diabetes Type** | | | |
| | Type 1 | 611 (1.5%) | 344 (2.8%) | 55 (2.3%) |
| | Type 2 | 5212 (13.1%) | 2932 (23.5%) | 581 (23.8%) |
| | **Hypertension** | 9549 (24%) | 5611 (45%) | 1047 (42.9%) |
| | **Malignant neoplasm** | 3798 (9.6%) | 1394 (11.2%) | 295 (12.1%) |
| | **Malnutrition** | 917 (2.3%) | 356 (2.9%) | NA |
| | **Mild to severe liver disease** | 1170 (2.9%) | 482 (3.9%) | 91 (3.7%) |
| | **Obesity** | 3399 (8.6%) | 1060 (8.5%) | 223 (9.1%) |
| | **Rheumatologic disorder** | 4273 (10.8%) | 1572 (12.6%) | 287 (11.8%) |
| | **Health worker** | 1737 (4.4%) | 561 (4.5%) | 62 (2.5%) |
| Severity of illness | **Asymptomatic** | 2262 (5.7%) | 2026 (16.2%) | 746 (30.5%) |
| | **Symptom onset (days)*** | | | |
| | Median (IQR) | 3 (IQR=7) | 2 (IQR=7) | 2 (IQR=7) |
| | Mean (SD) | 4.6 (SD=5.1) | 3.9 (SD=5.1) | 4 (SD=5) |
| | **Length of stay (days)** | | | |
| | Median (IQR) | 7 (IQR=9) | 8 (IQR=12) | 7 (IQR=11) |
| | Mean (SD) | 10.2 (SD=10.8) | 11.6 (SD=11.4) | 10.1 (SD=9.6) |
| | **ISARIC 4C Score** | | | |
| | Low (0-3) | 1844 (4.6%) | 665 (5.3%) | 179 (7.3%) |
| | Intermediate (4-8) | 5686 (14.3%) | 1697 (13.6%) | 417 (17.1%) |
| | High (9-14) | 13895 (35%) | 4940 (39.6%) | 921 (37.7%) |

| | | | | |
|---|---|---|---|---|
| | Very high (15+) | 4604 (11.6%) | 1239 (9.9%) | 138 (5.7%) |
| | Unknown | 13692 (34.5%) | 3928 (31.5%) | 787 (32.2%) |
| | **Respiratory rate (breaths/min)** | | | |
| | <20 | 11749 (29.6%) | 5018 (40.2%) | 1063 (43.5%) |
| | 20-30 | 20054 (50.5%) | 5592 (44.8%) | 1027 (42.1%) |
| | >=30 | 6055 (15.2%) | 1355 (10.9%) | 216 (8.8%) |
| | Unknown | 1863 (4.7%) | 504 (4%) | 136 (5.6%) |
| | **Peripheral oxygen saturation on room air (%)** | | | |
| | >=92 | 29794 (75%) | 10129 (81.2%) | 2038 (83.5%) |
| | <92 | 7953 (20%) | 1848 (14.8%) | 280 (11.5%) |
| | Unknown | 1974 (5%) | 492 (3.9%) | 124 (5.1%) |
| | **Glasgow coma score** | | | |
| | 15 | 30191 (76%) | 9884 (79.3%) | 2067 (84.6%) |
| | <15 | 5161 (13%) | 1700 (13.6%) | 205 (8.4%) |
| | Unknown | 4369 (11%) | 885 (7.1%) | 170 (7%) |
| | **Urea (mmol/L)** | | | |
| | <7 | 14265 (35.9%) | 4647 (37.3%) | 1014 (41.5%) |
| | 7-14 | 10675 (26.9%) | 3512 (28.2%) | 675 (27.6%) |
| | >14 | 5741 (14.5%) | 1707 (13.7%) | 256 (10.5%) |
| | Unknown | 9040 (22.8%) | 2603 (20.9%) | 497 (20.4%) |
| | **C-reactive protein (mg/dL)** | | | |
| | <50 | 10826 (27.3%) | 4496 (36.1%) | 947 (38.8%) |
| | 50-99 | 8131 (20.5%) | 2152 (17.3%) | 375 (15.4%) |
| | >=100 | 13510 (34%) | 3127 (25.1%) | 552 (22.6%) |
| | Unknown | 7254 (18.3%) | 2694 (21.6%) | 568 (23.3%) |
| **Respiratory support and treatments** | **Respiratory support** | | | |
| | None | 9314 (23.4%) | 4780 (38.3%) | 1184 (48.5%) |
| | Oxygen only | 27017 (68%) | 6910 (55.4%) | 1143 (46.8%) |
| | Non-invasive | 3390 (8.5%) | 779 (6.2%) | 115 (4.7%) |
| | **Steroids** | | | |
| | Yes | 3983 (10%) | 1463 (11.7%) | 637 (26.1%) |
| | No | 11079 (27.9%) | 6074 (48.7%) | 1623 (66.5%) |
| | Unknown | 24659 (62.1%) | 4932 (39.6%) | 182 (7.5%) |

* Symptom onset summary statistics based on patients with symptoms up to 3 weeks before admission only

Table E4: Baseline characteristics of ICU patients whose maximum respiratory support was oxygen only in the first wave, divided by 3 equal time points (N=1,344). Disclosure threshold of 5 was used.

| Type | Characteristic | 1 - Weeks 11 to 17<br>9 Mar to 26 Apr 2020 | 2 - Weeks 18 to 24<br>27 Apr to 14 Jun 2020 | 3 - Weeks 25 to 31<br>15 Jun to 2 Aug 2020 |
|---|---|---|---|---|
| | **Total** | 1006 (100%) | 260 (100%) | 78 (100%) |
| **Patient characteristics** | **Country** | | | |
| | England | 864 (85.9%) | 228 (87.7%) | 72 (92.3%) |
| | Scotland | 108 (10.7%) | 18 (6.9%) | NA |
| | Wales | 34 (3.4%) | 14 (5.4%) | 6 (7.7%) |
| | **Age (grouped)** | | | |
| | <50 | 200 (19.9%) | 59 (22.7%) | 20 (25.6%) |
| | 50-69 | 405 (40.3%) | 93 (35.8%) | 29 (37.2%) |
| | 70-79 | 217 (21.6%) | 40 (15.4%) | 29 (37.2%) |
| | 80+ | 184 (18.3%) | 68 (26.2%) | |
| | **Age (continuous)** | | | |
| | Median (IQR) | 64.1 (IQR=24) | 66.3 (IQR=29.1) | 64.6 (IQR=24.1) |
| | Mean (SD) | 64.1 (IQR=16.3) | 64.6 (IQR=18.4) | 61.4 (IQR=18) |
| | **Sex** | | | |
| | Female | 361 (35.9%) | 115 (44.2%) | 37 (47.4%) |
| | Male | 645 (64.1%) | 145 (55.8%) | 41 (52.6%) |
| | **Ethnic group** | | | |
| | White | 701 (69.7%) | 204 (78.5%) | 51 (65.4%) |
| | South Asian | 56 (5.6%) | 9 (3.5%) | 10 (12.8%) |
| | East Asian | 12 (1.2%) | NA | NA |
| | Black | 47 (4.7%) | 8 (3.1%) | NA |
| | Other Ethnic Minority | 64 (6.4%) | 21 (8.1%) | 11 (14.1%) |
| | Unknown | 126 (12.5%) | 18 (6.9%) | 6 (7.7%) |
| | **Number of comorbidities** | | | |
| | 0 | 289 (28.7%) | 48 (18.5%) | 16 (20.5%) |
| | 1 | 271 (26.9%) | 62 (23.8%) | 18 (23.1%) |
| | 2+ | 446 (44.3%) | 150 (57.7%) | 44 (56.4%) |
| | **AIDS/HIV** | 9 (0.9%) | NA | NA |
| | **Asthma** | 155 (15.4%) | 28 (10.8%) | 10 (12.8%) |
| | **Chronic cardiac disease** | 220 (21.9%) | 69 (26.5%) | 12 (15.4%) |
| | **Chronic hematologic disease** | 38 (3.8%) | 11 (4.2%) | NA |
| | **Chronic kidney disease** | 131 (13%) | 34 (13.1%) | 6 (7.7%) |
| | **Chronic neurological disorder** | 81 (8.1%) | 24 (9.2%) | 9 (11.5%) |
| | **Chronic pulmonary disease** | 117 (11.6%) | 36 (13.8%) | 13 (16.7%) |
| | **Dementia** | 70 (7%) | 27 (10.4%) | NA |
| | **Diabetes Type** | | | |
| | Type 1 | 19 (1.9%) | 10 (3.8%) | NA |
| | Type 2 | 117 (11.6%) | 63 (24.2%) | 11 (14.1%) |
| | **Hypertension** | 196 (19.5%) | 102 (39.2%) | 26 (33.3%) |
| | **Malignant neoplasm** | 66 (6.6%) | 34 (13.1%) | NA |
| | **Malnutrition** | 17 (1.7%) | 9 (3.5%) | NA |
| | **Mild to severe liver disease** | 28 (2.8%) | 13 (5%) | NA |
| | **Obesity** | 136 (13.5%) | 30 (11.5%) | 14 (17.9%) |
| | **Rheumatologic disorder** | 87 (8.6%) | 22 (8.5%) | 8 (10.3%) |
| | **Health worker** | 70 (7%) | 19 (7.3%) | NA |
| **Severity of illness** | **Asymptomatic** | 25 (2.5%) | 29 (11.2%) | 18 (23.1%) |
| | **Symptom onset (days)*** | | | |
| | Median (IQR) | 5 (IQR=8) | 3 (IQR=7) | 3 (IQR=6) |
| | Mean (SD) | 5.8 (IQR=5.3) | 4.6 (IQR=5.7) | 3.9 (IQR=4.9) |
| | **Length of stay (days)** | | | |
| | Median (IQR) | 9 (IQR=10) | 11 (IQR=13) | 9 (IQR=10.5) |
| | Mean (SD) | 12.2 (IQR=10.8) | 14.2 (IQR=11.8) | 11.4 (IQR=7.9) |
| | **ISARIC 4C Score** | | | |
| | Low to intermediate (0-8) | 286 (28.4%) | 78 (30%) | 25 (32.1%) |
| | High to very high (9+) | 371 (36.9%) | 99 (38.1%) | 26 (33.3%) |
| | Unknown | 349 (34.7%) | 83 (31.9%) | 27 (34.6%) |

| | | | | |
|---|---|---|---|---|
| | **Respiratory rate (breaths/min)** | | | |
| | <20 | 276 (27.4%) | 90 (34.6%) | 25 (32.1%) |
| | 20-30 | 514 (51.1%) | 130 (50%) | 39 (50%) |
| | >=30 | 182 (18.1%) | 35 (13.5%) | 13 (16.7%) |
| | Unknown | 34 (3.4%) | 5 (1.9%) | 1 (1.3%) |
| | **Peripheral oxygen saturation on room air (%)** | | | |
| | >=92 | 748 (74.4%) | 209 (80.4%) | 65 (83.3%) |
| | <92 | 214 (21.3%) | 46 (17.7%) | 12 (15.4%) |
| | Unknown | 44 (4.4%) | 5 (1.9%) | 1 (1.3%) |
| | **Glasgow coma score** | | | |
| | 15 | 788 (78.3%) | 199 (76.5%) | 64 (82.1%) |
| | <15 | 97 (9.6%) | 38 (14.6%) | 8 (10.3%) |
| | Unknown | 121 (12%) | 23 (8.8%) | 6 (7.7%) |
| | **Urea (mmol/L)** | | | |
| | <7 | 422 (41.9%) | 114 (43.8%) | 37 (47.4%) |
| | 7-14 | 242 (24.1%) | 62 (23.8%) | 17 (21.8%) |
| | >14 | 129 (12.8%) | 37 (14.2%) | 5 (6.4%) |
| | Unknown | 213 (21.2%) | 47 (18.1%) | 19 (24.4%) |
| | **C-reactive protein (mg/dL)** | | | |
| | <50 | 229 (22.8%) | 89 (34.2%) | 20 (25.6%) |
| | 50-99 | 199 (19.8%) | 45 (17.3%) | 18 (23.1%) |
| | >=100 | 431 (42.8%) | 86 (33.1%) | 23 (29.5%) |
| | Unknown | 147 (14.6%) | 40 (15.4%) | 17 (21.8%) |
| | **Steroids** | | | |
| | Yes | 123 (12.2%) | 41 (15.8%) | 40 (51.3%) |
| | No | 235 (23.4%) | 105 (40.4%) | 33 (42.3%) |
| | Unknown | 648 (64.4%) | 114 (43.8%) | 5 (6.4%) |

* Symptom onset summary statistics based on patients with symptoms up to 3 weeks before admission only

Table E5: Baseline characteristics of ICU patients whose maximum respiratory support was non-invasive ventilation only (including oxygen) in the first wave, divided by 3 equal time points (N=2,386). Disclosure threshold of 5 was used.

| Type | Characteristic | 1 - Weeks 11 to 17 9 Mar to 26 Apr 2020 | 2 - Weeks 18 to 24 27 Apr to 14 Jun 2020 | 3 - Weeks 25 to 31 15 Jun to 2 Aug 2020 |
|---|---|---|---|---|
| | **Total** | 1768 (100%) | 461 (100%) | 157 (100%) |
| Patient characteristics | **Country** | | | |
| | England | 1629 (92.1%) | 430 (93.3%) | 148 (94.3%) |
| | Scotland | 92 (5.2%) | 13 (2.8%) | NA |
| | Wales | 47 (2.7%) | 18 (3.9%) | 9 (5.7%) |
| | **Age (grouped)** | | | |
| | <50 | 362 (20.5%) | 85 (18.4%) | 26 (16.6%) |
| | 50-69 | 837 (47.3%) | 218 (47.3%) | 67 (42.7%) |
| | 70-79 | 406 (23%) | 116 (25.2%) | 64 (40.8%) |
| | 80+ | 163 (9.2%) | 42 (9.1%) | |
| | **Age (continuous)** | | | |
| | Median (IQR) | 62 (IQR=20.7) | 63.7 (IQR=20.9) | 65.9 (IQR=20.3) |
| | Mean (SD) | 61.6 (IQR=14.3) | 62.4 (IQR=14.6) | 65 (IQR=15.3) |
| | **Sex** | | | |
| | Female | 581 (32.9%) | 165 (35.8%) | 60 (38.2%) |
| | Male | 1187 (67.1%) | 296 (64.2%) | 97 (61.8%) |
| | **Ethnic group** | | | |
| | White | 1211 (68.5%) | 348 (75.5%) | 113 (72%) |
| | South Asian | 134 (7.6%) | 20 (4.3%) | 19 (12.1%) |
| | East Asian | 17 (1%) | NA | NA |
| | Black | 76 (4.3%) | 17 (3.7%) | NA |
| | Other Ethnic Minority | 161 (9.1%) | 29 (6.3%) | 18 (11.4%) |
| | Unknown | 169 (9.6%) | 47 (10.2%) | 7 (4.5%) |
| | **Number of comorbidities** | | | |
| | 0 | 522 (29.5%) | 80 (17.4%) | 19 (12.1%) |
| | 1 | 482 (27.3%) | 120 (26%) | 40 (25.5%) |
| | 2+ | 764 (43.2%) | 261 (56.6%) | 98 (62.4%) |
| | **AIDS/HIV** | 12 (0.7%) | NA | NA |
| | **Asthma** | 337 (19.1%) | 77 (16.7%) | 31 (19.7%) |
| | **Chronic cardiac disease** | 321 (18.2%) | 91 (19.7%) | 45 (28.7%) |
| | **Chronic hematologic disease** | 65 (3.7%) | 12 (2.6%) | NA |
| | **Chronic kidney disease** | 162 (9.2%) | 50 (10.8%) | 20 (12.7%) |
| | **Chronic neurological disorder** | 102 (5.8%) | 33 (7.2%) | 7 (4.5%) |
| | **Chronic pulmonary disease** | 252 (14.3%) | 89 (19.3%) | 39 (24.8%) |
| | **Dementia** | 24 (1.4%) | 8 (1.7%) | NA |
| | **Diabetes Type** | | | |
| | Type 1 | 31 (1.8%) | 13 (2.8%) | NA |
| | Type 2 | 252 (14.3%) | 132 (28.6%) | 38 (24.2%) |
| | **Hypertension** | 388 (21.9%) | 194 (42.1%) | 73 (46.5%) |
| | **Malignant neoplasm** | 91 (5.1%) | 40 (8.7%) | NA |
| | **Malnutrition** | 20 (1.1%) | 5 (1.1%) | NA |
| | **Mild to severe liver disease** | 40 (2.3%) | 13 (2.8%) | NA |
| | **Obesity** | 351 (19.9%) | 110 (23.9%) | 36 (22.9%) |
| | **Rheumatologic disorder** | 150 (8.5%) | 41 (8.9%) | 6 (3.8%) |
| | **Health worker** | 176 (10%) | 74 (16.1%) | NA |
| Severity of illness | **Asymptomatic** | 13 (0.7%) | 15 (3.3%) | 6 (3.8%) |
| | **Symptom onset (days)*** | | | |
| | Median (IQR) | 7 (IQR=7) | 5 (IQR=6.5) | 5 (IQR=6) |
| | Mean (SD) | 7 (IQR=4.9) | 5.3 (IQR=4.8) | 5.3 (IQR=4.5) |
| | **Length of stay (days)** | | | |
| | Median (IQR) | 11 (IQR=9) | 11 (IQR=12) | 10 (IQR=8) |
| | Mean (SD) | 12.9 (IQR=10) | 14.7 (IQR=12.7) | 11.7 (IQR=7.9) |
| | **ISARIC 4C Score** | | | |
| | Low to intermediate (0-8) | 514 (29.1%) | 142 (30.8%) | 49 (31.2%) |

| | | | | |
|---|---|---|---|---|
| | High to very high (9+) | 735 (41.6%) | 229 (49.7%) | 80 (51%) |
| | Unknown | 519 (29.4%) | 90 (19.5%) | 28 (17.8%) |
| | **Respiratory rate (breaths/min)** | | | |
| | <20 | 264 (14.9%) | 81 (17.6%) | 26 (16.6%) |
| | 20-30 | 924 (52.3%) | 267 (57.9%) | 86 (54.8%) |
| | >=30 | 529 (29.9%) | 110 (23.9%) | 44 (28%) |
| | Unknown | 51 (2.9%) | 3 (0.7%) | 1 (0.6%) |
| | **Peripheral oxygen saturation on room air (%)** | | | |
| | >=92 | 1078 (61%) | 294 (63.8%) | 101 (64.3%) |
| | <92 | 620 (35.1%) | 162 (35.1%) | 53 (33.8%) |
| | Unknown | 70 (4%) | 5 (1.1%) | 3 (1.9%) |
| | **Glasgow coma score** | | | |
| | 15 | 1498 (84.7%) | 405 (87.9%) | 132 (84.1%) |
| | <15 | 99 (5.6%) | 31 (6.7%) | 13 (8.3%) |
| | Unknown | 171 (9.7%) | 25 (5.4%) | 12 (7.6%) |
| | **Urea (mmol/L)** | | | |
| | <7 | 790 (44.7%) | 226 (49%) | 94 (59.9%) |
| | 7-14 | 464 (26.2%) | 127 (27.5%) | 36 (22.9%) |
| | >14 | 187 (10.6%) | 54 (11.7%) | 16 (10.2%) |
| | Unknown | 327 (18.5%) | 54 (11.7%) | 11 (7%) |
| | **C-reactive protein (mg/dL)** | | | |
| | <50 | 193 (10.9%) | 101 (21.9%) | 50 (31.8%) |
| | 50-99 | 361 (20.4%) | 93 (20.2%) | 24 (15.3%) |
| | >=100 | 977 (55.3%) | 229 (49.7%) | 68 (43.3%) |
| | Unknown | 237 (13.4%) | 38 (8.2%) | 15 (9.6%) |
| | **Steroids** | | | |
| | Yes | 270 (15.3%) | 148 (32.1%) | 110 (70.1%) |
| | No | 436 (24.7%) | 138 (29.9%) | 34 (21.7%) |
| | Unknown | 1062 (60.1%) | 175 (38%) | 13 (8.3%) |
| * Symptom onset summary statistics based on patients with symptoms up to 3 weeks before admission only | | | | |

Table E6: Baseline characteristics of ICU patients whose maximum respiratory support was invasive ventilation only (including oxygen) in the first wave, divided by 3 equal time points (N=5,610). Disclosure threshold of 5 was used.

| Type | Characteristic | 1 - Weeks 11 to 17<br>9 Mar to 26 Apr 2020 | 2 - Weeks 18 to 24<br>27 Apr to 14 Jun 2020 | 3 - Weeks 25 to 31<br>15 Jun to 2 Aug 2020 |
|---|---|---|---|---|
| | **Total** | 4958 (100%) | 554 (100%) | 98 (100%) |
| Patient characteristics | **Country** | | | |
| | England | 4480 (90.4%) | 495 (89.4%) | 92 (93.9%) |
| | Scotland | 260 (5.2%) | 22 (4%) | NA |
| | Wales | 218 (4.4%) | 37 (6.7%) | 6 (6.1%) |
| | **Age (grouped)** | | | |
| | <50 | 1002 (20.2%) | 122 (22%) | 33 (33.7%) |
| | 50-69 | 2952 (59.5%) | 326 (58.8%) | 50 (51%) |
| | 70-79 | 876 (17.7%) | 88 (15.9%) | 15 (15.3%) |
| | 80+ | 128 (2.6%) | 18 (3.2%) | |
| | **Age (continuous)** | | | |
| | Median (IQR) | 60.4 (IQR=15.8) | 59.6 (IQR=16.2) | 58.6 (IQR=19.8) |
| | Mean (SD) | 59.5 (IQR=12.2) | 58.8 (IQR=13) | 56.7 (IQR=14.4) |
| | **Sex** | | | |
| | Female | 1372 (27.7%) | 185 (33.4%) | 31 (31.6%) |
| | Male | 3586 (72.3%) | 369 (66.6%) | 67 (68.4%) |
| | **Ethnic group** | | | |
| | White | 2842 (57.3%) | 317 (57.2%) | 45 (45.9%) |
| | South Asian | 372 (7.5%) | 58 (10.5%) | 25 (25.5%) |
| | East Asian | 95 (1.9%) | NA | NA |
| | Black | 345 (7%) | 32 (5.8%) | NA |
| | Other Ethnic Minority | 606 (12.2%) | 72 (13%) | 19 (19.4%) |
| | Unknown | 698 (14.1%) | 75 (13.5%) | 9 (9.2%) |
| | **Number of comorbidities** | | | |
| | 0 | 1844 (37.2%) | 131 (23.6%) | 22 (22.4%) |
| | 1 | 1485 (30%) | 136 (24.5%) | 22 (22.4%) |
| | 2+ | 1629 (32.9%) | 287 (51.8%) | 54 (55.1%) |
| | **AIDS/HIV** | 33 (0.7%) | NA | NA |
| | **Asthma** | 750 (15.1%) | 82 (14.8%) | 13 (13.3%) |
| | **Chronic cardiac disease** | 606 (12.2%) | 90 (16.2%) | 19 (19.4%) |
| | **Chronic hematologic disease** | 119 (2.4%) | 11 (2%) | NA |
| | **Chronic kidney disease** | 279 (5.6%) | 45 (8.1%) | 9 (9.2%) |
| | **Chronic neurological disorder** | 192 (3.9%) | 30 (5.4%) | 9 (9.2%) |
| | **Chronic pulmonary disease** | 324 (6.5%) | 47 (8.5%) | 9 (9.2%) |
| | **Dementia** | 47 (0.9%) | 10 (1.8%) | NA |
| | **Diabetes Type** | | | |
| | Type 1 | 59 (1.2%) | 26 (4.7%) | NA |
| | Type 2 | 585 (11.8%) | 148 (26.7%) | 29 (29.6%) |
| | **Hypertension** | 928 (18.7%) | 226 (40.8%) | 39 (39.8%) |
| | **Malignant neoplasm** | 212 (4.3%) | 27 (4.9%) | NA |
| | **Malnutrition** | 34 (0.7%) | 5 (0.9%) | NA |
| | **Mild to severe liver disease** | 104 (2.1%) | 19 (3.4%) | NA |
| | **Obesity** | 1037 (20.9%) | 142 (25.6%) | 21 (21.4%) |
| | **Rheumatologic disorder** | 294 (5.9%) | 39 (7%) | 7 (7.1%) |
| | **Health worker** | 436 (8.8%) | 84 (15.2%) | NA |
| Severity of illness | **Asymptomatic** | 40 (0.8%) | 30 (5.4%) | 13 (13.3%) |
| | **Symptom onset (days)*** | | | |
| | Median (IQR) | 7 (IQR=6) | 6 (IQR=6) | 4 (IQR=5) |
| | Mean (SD) | 7.2 (IQR=4.7) | 6.1 (IQR=4.9) | 5 (IQR=4.4) |
| | **Length of stay (days)** | | | |
| | Median (IQR) | 19 (IQR=21) | 19 (IQR=18) | 18.5 (IQR=15.8) |
| | Mean (SD) | 23.6 (IQR=19.4) | 22.3 (IQR=16.6) | 19.2 (IQR=12.6) |
| | **ISARIC 4C Score** | | | |
| | Low to intermediate (0-8) | 1207 (24.3%) | 166 (30%) | 33 (33.7%) |

| | | | | |
|---|---|---|---|---|
| | High to very high (9+) | 2027 (40.9%) | 242 (43.7%) | 35 (35.7%) |
| | Unknown | 1724 (34.8%) | 146 (26.4%) | 30 (30.6%) |
| | **Respiratory rate (breaths/min)** | | | |
| | <20 | 777 (15.7%) | 114 (20.6%) | 17 (17.3%) |
| | 20-30 | 2299 (46.4%) | 243 (43.9%) | 45 (45.9%) |
| | >=30 | 1668 (33.6%) | 170 (30.7%) | 32 (32.7%) |
| | Unknown | 214 (4.3%) | 27 (4.9%) | 4 (4.1%) |
| | **Peripheral oxygen saturation on room air (%)** | | | |
| | >=92 | 2726 (55%) | 338 (61%) | 55 (56.1%) |
| | <92 | 2020 (40.7%) | 195 (35.2%) | 39 (39.8%) |
| | Unknown | 212 (4.3%) | 21 (3.8%) | 4 (4.1%) |
| | **Glasgow coma score** | | | |
| | 15 | 2926 (59%) | 369 (66.6%) | 60 (61.2%) |
| | <15 | 1288 (26%) | 142 (25.6%) | 24 (24.5%) |
| | Unknown | 744 (15%) | 43 (7.8%) | 14 (14.3%) |
| | **Urea (mmol/L)** | | | |
| | <7 | 2104 (42.4%) | 263 (47.5%) | 50 (51%) |
| | 7-14 | 1404 (28.3%) | 157 (28.3%) | 29 (29.6%) |
| | >14 | 515 (10.4%) | 71 (12.8%) | 9 (9.2%) |
| | Unknown | 935 (18.9%) | 63 (11.4%) | 10 (10.2%) |
| | **C-reactive protein (mg/dL)** | | | |
| | <50 | 466 (9.4%) | 89 (16.1%) | 20 (20.4%) |
| | 50-99 | 709 (14.3%) | 107 (19.3%) | 25 (25.5%) |
| | >=100 | 3030 (61.1%) | 294 (53.1%) | 41 (41.8%) |
| | Unknown | 753 (15.2%) | 64 (11.6%) | 12 (12.2%) |
| | **Steroids** | | | |
| | Yes | 1274 (25.7%) | 189 (34.1%) | 67 (68.4%) |
| | No | 1108 (22.3%) | 190 (34.3%) | 22 (22.4%) |
| | Unknown | 2576 (52%) | 175 (31.6%) | 9 (9.2%) |
| * Symptom onset summary statistics based on patients with symptoms up to 3 weeks before admission only | | | | |

Table E7: Baseline characteristics of ward patients who received no respiratory support in the first wave, divided by 3 equal time points (N=15,278). Disclosure threshold of 5 was used.

| Type | Characteristic | 1 - Weeks 11 to 17 9 Mar to 26 Apr 2020 | 2 - Weeks 18 to 24 27 Apr to 14 Jun 2020 | 3 - Weeks 25 to 31 15 Jun to 2 Aug 2020 |
|---|---|---|---|---|
| | **Total** | 9314 (100%) | 4780 (100%) | 1184 (100%) |
| Patient characteristics | **Country** | | | |
| | England | 8324 (89.4%) | 4363 (91.3%) | 1091 (92.1%) |
| | Scotland | 515 (5.5%) | 210 (4.4%) | 19 (1.6%) |
| | Wales | 475 (5.1%) | 207 (4.3%) | 74 (6.2%) |
| | **Age (grouped)** | | | |
| | <50 | 1700 (18.3%) | 791 (16.5%) | 242 (20.4%) |
| | 50-69 | 2415 (25.9%) | 974 (20.4%) | 287 (24.2%) |
| | 70-79 | 1880 (20.2%) | 986 (20.6%) | 214 (18.1%) |
| | 80+ | 3319 (35.6%) | 2029 (42.4%) | 441 (37.2%) |
| | **Age (continuous)** | | | |
| | Median (IQR) | 73.1 (IQR=28.4) | 76.6 (IQR=26.4) | 73.6 (IQR=28.9) |
| | Mean (SD) | 68.7 (IQR=19) | 71.2 (IQR=19.4) | 68 (IQR=20.3) |
| | **Sex** | | | |
| | Female | 4474 (48%) | 2523 (52.8%) | 614 (51.9%) |
| | Male | 4840 (52%) | 2257 (47.2%) | 570 (48.1%) |
| | **Ethnic group** | | | |
| | White | 6530 (70.1%) | 3762 (78.7%) | 828 (69.9%) |
| | South Asian | 515 (5.5%) | 215 (4.5%) | 117 (9.9%) |
| | East Asian | 70 (0.8%) | NA | NA |
| | Black | 403 (4.3%) | 87 (1.8%) | NA |
| | Other Ethnic Minority | 600 (6.4%) | 237 (5%) | 112 (9.5%) |
| | Unknown | 1196 (12.8%) | 479 (10%) | 127 (10.7%) |
| | **Number of comorbidities** | | | |
| | 0 | 2667 (28.6%) | 1006 (21%) | 298 (25.2%) |
| | 1 | 2018 (21.7%) | 848 (17.7%) | 225 (19%) |
| | 2+ | 4629 (49.7%) | 2926 (61.2%) | 661 (55.8%) |
| | **AIDS/HIV** | 29 (0.3%) | 12 (0.3%) | NA |
| | **Asthma** | 1125 (12.1%) | 536 (11.2%) | 121 (10.2%) |
| | **Chronic cardiac disease** | 2492 (26.8%) | 1496 (31.3%) | 355 (30%) |
| | **Chronic hematologic disease** | 335 (3.6%) | 200 (4.2%) | 45 (3.8%) |
| | **Chronic kidney disease** | 1356 (14.6%) | 823 (17.2%) | 213 (18%) |
| | **Chronic neurological disorder** | 973 (10.4%) | 546 (11.4%) | 122 (10.3%) |
| | **Chronic pulmonary disease** | 1171 (12.6%) | 623 (13%) | 134 (11.3%) |
| | **Dementia** | 1436 (15.4%) | 907 (19%) | 161 (13.6%) |
| | **Diabetes Type** | | | |
| | Type 1 | 187 (2%) | 142 (3%) | NA |
| | Type 2 | 1257 (13.5%) | 991 (20.7%) | 256 (21.6%) |
| | **Hypertension** | 2303 (24.7%) | 1996 (41.8%) | 461 (38.9%) |
| | **Malignant neoplasm** | 774 (8.3%) | 479 (10%) | 134 (11.3%) |
| | **Malnutrition** | 215 (2.3%) | 130 (2.7%) | NA |
| | **Mild to severe liver disease** | 304 (3.3%) | 186 (3.9%) | 44 (3.7%) |
| | **Obesity** | 601 (6.5%) | 339 (7.1%) | 81 (6.8%) |
| | **Rheumatologic disorder** | 900 (9.7%) | 545 (11.4%) | 123 (10.4%) |
| | **Health worker** | 601 (6.5%) | 259 (5.4%) | NA |
| Severity of illness | **Asymptomatic** | 656 (7%) | 1069 (22.4%) | 446 (37.7%) |
| | **Symptom onset (days)*** | | | |
| | Median (IQR) | 2 (IQR=7) | 1 (IQR=6) | 2 (IQR=6) |
| | Mean (SD) | 4 (IQR=5.2) | 3.4 (IQR=5.1) | 3.7 (IQR=4.9) |
| | **Length of stay (days)** | | | |
| | Median (IQR) | 6 (IQR=11) | 7 (IQR=13) | 6 (IQR=11) |
| | Mean (SD) | 9.9 (IQR=12.5) | 10.9 (IQR=12.1) | 9.6 (IQR=10.1) |
| | **ISARIC 4C Score** | | | |
| | Low (0-3) | 732 (7.9%) | 391 (8.2%) | 110 (9.3%) |
| | Intermediate (4-8) | 1288 (13.8%) | 662 (13.8%) | 196 (16.6%) |
| | High (9-14) | 2460 (26.4%) | 1626 (34%) | 356 (30.1%) |

| | | | | |
|---|---|---|---|---|
| | Very high (15+) | 333 (3.6%) | 150 (3.1%) | 23 (1.9%) |
| | Unknown | 4501 (48.3%) | 1951 (40.8%) | 499 (42.1%) |
| | **Respiratory rate (breaths/min)** | | | |
| | <20 | 3501 (37.6%) | 1624 (34%) | 377 (31.8%) |
| | 20-30 | 604 (6.5%) | 180 (3.8%) | 50 (4.2%) |
| | >=30 | 1268 (13.6%) | 423 (8.8%) | 117 (9.9%) |
| | Unknown | 3501 (37.6%) | 1624 (34%) | 377 (31.8%) |
| | **Peripheral oxygen saturation on room air (%)** | | | |
| | >=92 | 7497 (80.5%) | 4200 (87.9%) | 1044 (88.2%) |
| | <92 | 496 (5.3%) | 152 (3.2%) | 29 (2.4%) |
| | Unknown | 1321 (14.2%) | 428 (9%) | 111 (9.4%) |
| | **Glasgow coma score** | | | |
| | 15 | 6491 (69.7%) | 3769 (78.8%) | 978 (82.6%) |
| | <15 | 758 (8.1%) | 467 (9.8%) | 66 (5.6%) |
| | Unknown | 2065 (22.2%) | 544 (11.4%) | 140 (11.8%) |
| | **Urea (mmol/L)** | | | |
| | <7 | 3449 (37%) | 1865 (39%) | 496 (41.9%) |
| | 7-14 | 1915 (20.6%) | 1082 (22.6%) | 257 (21.7%) |
| | >14 | 850 (9.1%) | 467 (9.8%) | 104 (8.8%) |
| | Unknown | 3100 (33.3%) | 1366 (28.6%) | 327 (27.6%) |
| | **C-reactive protein (mg/dL)** | | | |
| | <50 | 3406 (36.6%) | 2053 (42.9%) | 509 (43%) |
| | 50-99 | 1493 (16%) | 622 (13%) | 130 (11%) |
| | >=100 | 1709 (18.3%) | 660 (13.8%) | 171 (14.4%) |
| | Unknown | 2706 (29.1%) | 1445 (30.2%) | 374 (31.6%) |
| | **Steroids** | | | |
| | Yes | 580 (6.2%) | 332 (6.9%) | 119 (10.1%) |
| | No | 2782 (29.9%) | 2468 (51.6%) | 947 (80%) |
| | Unknown | 5952 (63.9%) | 1980 (41.4%) | 118 (10%) |
| * Symptom onset summary statistics based on patients with symptoms up to 3 weeks before admission only | | | | |

Table E8: Baseline characteristics of ward patients whose maximum respiratory support oxygen only in the first wave, divided by 3 equal time points (N=35,070). Disclosure threshold of 5 was used.

| Type | Characteristic | 1 - Weeks 11 to 17<br>9 Mar to 26 Apr 2020 | 2 - Weeks 18 to 24<br>27 Apr to 14 Jun 2020 | 3 - Weeks 25 to 31<br>15 Jun to 2 Aug 2020 |
|---|---|---|---|---|
| | **Total** | 27017 (100%) | 6910 (100%) | 1143 (100%) |
| Patient characteristics | **Country** | | | |
| | England | 24870 (92.1%) | 6333 (91.6%) | 1081 (94.6%) |
| | Scotland | 1116 (4.1%) | 267 (3.9%) | 13 (1.1%) |
| | Wales | 1031 (3.8%) | 310 (4.5%) | 49 (4.3%) |
| | Age (grouped) | | | |
| | <50 | 2717 (10.1%) | 540 (7.8%) | 155 (13.6%) |
| | 50-69 | 6620 (24.5%) | 1432 (20.7%) | 271 (23.7%) |
| | 70-79 | 6343 (23.5%) | 1566 (22.7%) | 265 (23.2%) |
| | 80+ | 11337 (42%) | 3372 (48.8%) | 452 (39.5%) |
| | Age (continuous) | | | |
| | Median (IQR) | 77 (IQR=22) | 79.5 (IQR=19.4) | 76.3 (IQR=23.7) |
| | Mean (SD) | 73.1 (IQR=15.9) | 75.5 (IQR=15.4) | 71.9 (IQR=17.1) |
| | Sex | | | |
| | Female | 11762 (43.5%) | 3335 (48.3%) | 552 (48.3%) |
| | Male | 15255 (56.5%) | 3575 (51.7%) | 591 (51.7%) |
| | Ethnic group | | | |
| | White | 20233 (74.9%) | 5604 (81.1%) | 802 (70.2%) |
| | South Asian | 1053 (3.9%) | 183 (2.6%) | 107 (9.4%) |
| | East Asian | 190 (0.7%) | NA | NA |
| | Black | 1007 (3.7%) | 81 (1.2%) | NA |
| | Other Ethnic Minority | 1650 (6.1%) | 322 (4.6%) | 114 (9.9%) |
| | Unknown | 2884 (10.7%) | 720 (10.4%) | 120 (10.5%) |
| | Number of comorbidities | | | |
| | 0 | 4824 (17.9%) | 688 (10%) | 141 (12.3%) |
| | 1 | 6305 (23.3%) | 1161 (16.8%) | 219 (19.2%) |
| | 2+ | 15888 (58.8%) | 5061 (73.2%) | 783 (68.5%) |
| | **AIDS/HIV** | 80 (0.3%) | 20 (0.3%) | NA |
| | **Asthma** | 3515 (13%) | 783 (11.3%) | 145 (12.7%) |
| | **Chronic cardiac disease** | 9116 (33.7%) | 2698 (39%) | 407 (35.6%) |
| | **Chronic hematologic disease** | 1124 (4.2%) | 366 (5.3%) | 57 (5%) |
| | **Chronic kidney disease** | 5016 (18.6%) | 1427 (20.7%) | 213 (18.6%) |
| | **Chronic neurological disorder** | 3636 (13.5%) | 1096 (15.9%) | 145 (12.7%) |
| | **Chronic pulmonary disease** | 5124 (19%) | 1467 (21.2%) | 251 (22%) |
| | **Dementia** | 5079 (18.8%) | 1604 (23.2%) | 169 (14.8%) |
| | **Diabetes Type** | | | |
| | Type 1 | 387 (1.4%) | 181 (2.6%) | NA |
| | Type 2 | 3477 (12.9%) | 1701 (24.6%) | 284 (24.8%) |
| | **Hypertension** | 6437 (23.8%) | 3217 (46.6%) | 530 (46.4%) |
| | **Malignant neoplasm** | 2751 (10.2%) | 834 (12.1%) | 148 (12.9%) |
| | **Malnutrition** | 639 (2.4%) | 209 (3%) | NA |
| | **Mild to severe liver disease** | 763 (2.8%) | 262 (3.8%) | 42 (3.7%) |
| | **Obesity** | 2362 (8.7%) | 592 (8.6%) | 126 (11%) |
| | **Rheumatologic disorder** | 2993 (11.1%) | 925 (13.4%) | 152 (13.3%) |
| | **Health worker** | 1016 (3.8%) | 271 (3.9%) | NA |
| Severity of illness | **Asymptomatic** | 588 (2.2%) | 588 (8.5%) | 202 (17.7%) |
| | **Symptom onset (days)** | | | |
| | Median (IQR) | 3 (IQR=7) | 2 (IQR=7) | 3 (IQR=7) |
| | Mean (SD) | 4.7 (IQR=5.1) | 4.1 (IQR=5.1) | 4.1 (IQR=4.9) |
| | **Length of stay (days)** | | | |
| | Median (IQR) | 7 (IQR=9) | 9 (IQR=11) | 8 (IQR=11) |
| | Mean (SD) | 10.3 (IQR=10.3) | 12.1 (IQR=10.9) | 10.7 (IQR=9) |
| | **ISARIC 4C Score** | | | |
| | Low (0-3) | 1044 (3.9%) | 258 (3.7%) | 63 (5.5%) |
| | Intermediate (4-8) | 3891 (14.4%) | 914 (13.2%) | 210 (18.4%) |
| | High (9-14) | 10095 (37.4%) | 2958 (42.8%) | 497 (43.5%) |

| | | | | |
|---|---|---|---|---|
| | Very high (15+) | 3736 (13.8%) | 945 (13.7%) | 99 (8.7%) |
| | Unknown | 8251 (30.5%) | 1835 (26.6%) | 274 (24%) |
| | **Respiratory rate (breaths/min)** | | | |
| | <20 | 7194 (26.6%) | 2289 (33.1%) | 403 (35.3%) |
| | 20-30 | 14689 (54.4%) | 3543 (51.3%) | 587 (51.4%) |
| | >=30 | 4612 (17.1%) | 1003 (14.5%) | 135 (11.8%) |
| | Unknown | 522 (1.9%) | 75 (1.1%) | 18 (1.6%) |
| | **Peripheral oxygen saturation on room air (%)** | | | |
| | >=92 | 20109 (74.4%) | 5413 (78.3%) | 924 (80.8%) |
| | <92 | 6216 (23%) | 1413 (20.4%) | 201 (17.6%) |
| | Unknown | 692 (2.6%) | 84 (1.2%) | 18 (1.6%) |
| | **Glasgow coma score** | | | |
| | 15 | 21006 (77.8%) | 5482 (79.3%) | 991 (86.7%) |
| | <15 | 4014 (14.9%) | 1114 (16.1%) | 126 (11%) |
| | Unknown | 1997 (7.4%) | 314 (4.5%) | 26 (2.3%) |
| | **Urea (mmol/L)** | | | |
| | <7 | 9644 (35.7%) | 2508 (36.3%) | 476 (41.6%) |
| | 7-14 | 7724 (28.6%) | 2163 (31.3%) | 369 (32.3%) |
| | >14 | 4346 (16.1%) | 1091 (15.8%) | 139 (12.2%) |
| | Unknown | 5303 (19.6%) | 1148 (16.6%) | 159 (13.9%) |
| | **C-reactive protein (mg/dL)** | | | |
| | <50 | 6785 (25.1%) | 2242 (32.4%) | 402 (35.2%) |
| | 50-99 | 5929 (21.9%) | 1367 (19.8%) | 212 (18.5%) |
| | >=100 | 10203 (37.8%) | 2141 (31%) | 343 (30%) |
| | Unknown | 4100 (15.2%) | 1160 (16.8%) | 186 (16.3%) |
| **Respiratory support and treatments** | **Steroids** | | | |
| | Yes | 2883 (10.7%) | 957 (13.8%) | 442 (38.7%) |
| | No | 7514 (27.8%) | 3266 (47.3%) | 640 (56%) |
| | Unknown | 16620 (61.5%) | 2687 (38.9%) | 61 (5.3%) |

* Symptom onset summary statistics based on patients with symptoms up to 3 weeks before admission only

Table E9: Baseline characteristics of ward patients who received non-invasive ventilation only (including oxygen) in the first wave, divided by 3 equal time points (N=4,284). Disclosure threshold of 5 was used.

| Type | Characteristic | 1 - Weeks 11 to 17 9 Mar to 26 Apr 2020 | 2 - Weeks 18 to 24 27 Apr to 14 Jun 2020 | 3 - Weeks 25 to 31 15 Jun to 2 Aug 2020 |
|---|---|---|---|---|
| | **Total** | 3390 (100%) | 779 (100%) | 115 (100%) |
| Patient characteristics | **Country** | | | |
| | England | 3159 (93.2%) | 730 (93.7%) | 105 (91.3%) |
| | Scotland | 49 (1.4%) | 4 (0.5%) | 1 (0.9%) |
| | Wales | 182 (5.4%) | 45 (5.8%) | 9 (7.8%) |
| | **Age (grouped)** | | | |
| | <50 | 280 (8.3%) | 66 (8.5%) | 10 (8.7%) |
| | 50-69 | 1055 (31.1%) | 218 (28%) | 30 (26.1%) |
| | 70-79 | 953 (28.1%) | 219 (28.1%) | 42 (36.5%) |
| | 80+ | 1102 (32.5%) | 276 (35.4%) | 33 (28.7%) |
| | **Age (continuous)** | | | |
| | Median (IQR) | 74 (IQR=20.3) | 75.3 (IQR=19.4) | 74.2 (IQR=17.9) |
| | Mean (SD) | 71.5 (IQR=14.3) | 72.6 (IQR=14.2) | 71.8 (IQR=13.8) |
| | **Sex** | | | |
| | Male | 2103 (62%) | 443 (56.9%) | 56 (48.7%) |
| | Female | 1287 (38%) | 336 (43.1%) | 59 (51.3%) |
| | **Ethnic group** | | | |
| | White | 2476 (73%) | 597 (76.6%) | 83 (72.2%) |
| | South Asian | 149 (4.4%) | 45 (5.8%) | 18 (15.7%) |
| | East Asian | 14 (0.4%) | NA | NA |
| | Black | 137 (4%) | 11 (1.4%) | NA |
| | Other Ethnic Minority | 259 (7.6%) | 41 (5.2%) | 8 (6.9%) |
| | Unknown | 355 (10.5%) | 85 (10.9%) | 6 (5.2%) |
| | **Number of comorbidities** | | | |
| | 0 | 643 (19%) | 61 (7.8%) | 11 (9.6%) |
| | 1 | 828 (24.4%) | 136 (17.5%) | 22 (19.1%) |
| | 2+ | 1919 (56.6%) | 582 (74.7%) | 82 (71.3%) |
| | **AIDS/HIV** | 15 (0.4%) | NA | NA |
| | **Asthma** | 491 (14.5%) | 106 (13.6%) | 16 (13.9%) |
| | **Chronic cardiac disease** | 1082 (31.9%) | 301 (38.6%) | 45 (39.1%) |
| | **Chronic hematologic disease** | 152 (4.5%) | 33 (4.2%) | 6 (5.2%) |
| | **Chronic kidney disease** | 516 (15.2%) | 172 (22.1%) | 22 (19.1%) |
| | **Chronic neurological disorder** | 354 (10.4%) | 100 (12.8%) | 12 (10.4%) |
| | **Chronic pulmonary disease** | 785 (23.2%) | 220 (28.2%) | 36 (31.3%) |
| | **Dementia** | 336 (9.9%) | 110 (14.1%) | 8 (7%) |
| | **Diabetes Type** | | | |
| | Type 1 | 37 (1.1%) | 21 (2.7%) | NA |
| | Type 2 | 478 (14.1%) | 240 (30.8%) | 41 (35.7%) |
| | **Hypertension** | 809 (23.9%) | 398 (51.1%) | 56 (48.7%) |
| | **Malignant neoplasm** | 273 (8.1%) | 81 (10.4%) | 13 (11.3%) |
| | **Malnutrition** | 63 (1.9%) | 17 (2.2%) | NA |
| | **Mild to severe liver disease** | 103 (3%) | 34 (4.4%) | 5 (4.3%) |
| | **Obesity** | 436 (12.9%) | 129 (16.6%) | 16 (13.9%) |
| | **Rheumatologic disorder** | 380 (11.2%) | 102 (13.1%) | 12 (10.4%) |
| | **Health worker** | 120 (3.5%) | 31 (4%) | NA |
| Severity of illness | **Asymptomatic** | 49 (1.4%) | 35 (4.5%) | 7 (6.1%) |
| | **Symptom onset (days)*** | | | |
| | Median (IQR) | 4 (IQR=7) | 3 (IQR=7) | 3 (IQR=7) |
| | Mean (SD) | 5.3 (IQR=5.2) | 4.7 (IQR=5.4) | 5.1 (IQR=5.8) |
| | **Length of stay (days)** | | | |
| | Median (IQR) | 8 (IQR=9) | 8 (IQR=10) | 8 (IQR=10) |
| | Mean (SD) | 10.5 (IQR=9) | 11.9 (IQR=10.9) | 10.8 (IQR=10) |
| | **ISARIC 4C Score** | | | |
| | Low (0-3) | 65 (1.9%) | 15 (1.9%) | 6 (5.2%) |

| | | | | |
|---|---|---|---|---|
| | Intermediate (4-8) | 498 (14.7%) | 120 (15.4%) | 10 (8.7%) |
| | High (9-14) | 1305 (38.5%) | 350 (44.9%) | 67 (58.3%) |
| | Very high (15+) | 502 (14.8%) | 138 (17.7%) | 15 (13%) |
| | Unknown | 1020 (30.1%) | 156 (20%) | 17 (14.8%) |
| | **Respiratory rate (breaths/min)** | | | |
| | <20 | 614 (18.1%) | 176 (22.6%) | 20 (17.4%) |
| | 20-30 | 1864 (55%) | 425 (54.6%) | 63 (54.8%) |
| | >=30 | 838 (24.7%) | 172 (22.1%) | 31 (27%) |
| | Unknown | 74 (2.2%) | 6 (0.8%) | 1 (0.9%) |
| | **Peripheral oxygen saturation on room air (%)** | | | |
| | >=92 | 2187 (64.5%) | 516 (66.2%) | 70 (60.9%) |
| | <92 | 1101 (32.5%) | 256 (32.9%) | 44 (38.3%) |
| | Unknown | 102 (3%) | 7 (0.9%) | 1 (0.9%) |
| | **Glasgow coma score** | | | |
| | 15 | 2694 (79.5%) | 633 (81.3%) | 98 (85.2%) |
| | <15 | 389 (11.5%) | 119 (15.3%) | 13 (11.3%) |
| | Unknown | 307 (9.1%) | 27 (3.5%) | 4 (3.5%) |
| | **Urea (mmol/L)** | | | |
| | <7 | 1172 (34.6%) | 274 (35.2%) | 42 (36.5%) |
| | 7-14 | 1036 (30.6%) | 267 (34.3%) | 49 (42.6%) |
| | >14 | 545 (16.1%) | 149 (19.1%) | 13 (11.3%) |
| | Unknown | 637 (18.8%) | 89 (11.4%) | 11 (9.6%) |
| | **C-reactive protein (mg/dL)** | | | |
| | <50 | 635 (18.7%) | 201 (25.8%) | 36 (31.3%) |
| | 50-99 | 709 (20.9%) | 163 (20.9%) | 33 (28.7%) |
| | >=100 | 1598 (47.1%) | 326 (41.8%) | 38 (33%) |
| | Unknown | 448 (13.2%) | 89 (11.4%) | 8 (7%) |
| **Respiratory support and treatments** | **Steroids** | | | |
| | Yes | 520 (15.3%) | 174 (22.3%) | 76 (66.1%) |
| | No | 783 (23.1%) | 340 (43.6%) | 36 (31.3%) |
| | Unknown | 2087 (61.6%) | 265 (34%) | 3 (2.6%) |

* Symptom onset summary statistics based on patients with symptoms up to 3 weeks before admission only

Table E10: In-patient mortality and 95% CIs for 3 equal time-periods (Weeks 11 to 17, 18 to 24 and 25 to 31). 95% confidence intervals calculated by the Exact method.

| | | | 1 - Weeks 11 to 17 9 Mar to 26 Apr 2020 | | 2 - Weeks 18 to 24 27 Apr to 14 Jun 2020 | | 3 - Weeks 25 to 31 15 Jun to 2 Aug 2020 | |
|---|---|---|---|---|---|---|---|---|
| **Threshold of care** | **Respiratory support** | **Age group** | **Mortality (%)** | **95% CI** | **Mortality (%)** | **95% CI** | **Mortality (%)** | **95% CI** |
| **Overall** | | **Overall** | 32.3 | (31.8, 32.7) | 24.9 | (24.1, 25.6) | 16.4 | (15, 17.8) |
| | | <50 | 6.1 | (5.5, 6.7) | 3.6 | (2.8, 4.6) | 3.1 | (1.7, 5) |
| | | 50-69 | 20.5 | (19.9, 21.2) | 15 | (13.8, 16.2) | 10.6 | (8.5, 13.1) |
| | | 70-79 | 38.7 | (37.7, 39.6) | 28 | (26.4, 29.6) | 20.8 | (17.6, 24.4) |
| | | 80+ | 48.5 | (47.7, 49.2) | 34.9 | (33.6, 36.1) | 24.8 | (22.1, 27.6) |
| **ICU** | **Invasive** | **Overall** | 41 | (39.7, 42.4) | 37.7 | (33.7, 41.9) | 41.8 | (31.9, 52.2) |
| | | <50 | 20.6 | (18.1, 23.2) | 20.5 | (13.7, 28.7) | 21.2 | (9, 38.9) |
| | | 50-69 | 40 | (38.3, 41.8) | 35.6 | (30.4, 41) | 48 | (33.7, 62.6) |
| | | 70-79 | 63 | (59.7, 66.2) | 63.6 | (52.7, 73.6) | 69.2 | (38.6, 90.9) |
| | | 80+ | 73.4 | (64.9, 80.9) | 66.7 | (41, 86.7) | 50 | (1.3, 98.7) |
| | **Non-invasive** | **Overall** | 33.2 | (31, 35.5) | 25.6 | (21.7, 29.8) | 24.8 | (18.3, 32.4) |
| | | <50 | 7.7 | (5.2, 11) | 4.7 | (1.3, 11.6) | 11.5 | (2.4, 30.2) |
| | | 50-69 | 23.7 | (20.8, 26.7) | 16.1 | (11.4, 21.6) | 19.4 | (10.8, 30.9) |
| | | 70-79 | 56.4 | (51.4, 61.3) | 43.1 | (33.9, 52.6) | 43.2 | (27.1, 60.5) |
| | | 80+ | 81 | (74.1, 86.7) | 69 | (52.9, 82.4) | 25.9 | (11.1, 46.3) |
| | **Oxygen only** | **Overall** | 22.7 | (20.1, 25.4) | 19.6 | (15, 25) | 12.8 | (6.3, 22.3) |
| | | <50 | 2.5 | (0.8, 5.7) | 5.1 | (1.1, 14.1) | 0 | (0, 16.8) |
| | | 50-69 | 16.5 | (13.1, 20.5) | 14 | (7.7, 22.7) | 10.3 | (2.2, 27.4) |
| | | 70-79 | 35.5 | (29.1, 42.2) | 25 | (12.7, 41.2) | 21.1 | (6.1, 45.6) |
| | | 80+ | 42.9 | (35.7, 50.4) | 36.8 | (25.4, 49.3) | 30 | (6.7, 65.2) |
| **Ward** | **None** | **Overall** | 14.4 | (13.7, 15.1) | 8.4 | (7.7, 9.3) | 6 | (4.7, 7.5) |
| | | <50 | 1.2 | (0.7, 1.8) | 0.8 | (0.3, 1.6) | 0.4 | (0, 2.3) |
| | | 50-69 | 6.4 | (5.4, 7.4) | 4 | (2.9, 5.4) | 1.7 | (0.6, 4) |
| | | 70-79 | 16.6 | (15, 18.4) | 9.3 | (7.6, 11.3) | 5.6 | (2.9, 9.6) |
| | | 80+ | 25.7 | (24.2, 27.2) | 13.1 | (11.7, 14.7) | 12 | (9.1, 15.4) |
| | **Non-invasive** | **Overall** | 48 | (46.3, 49.7) | 49.9 | (46.4, 53.5) | 44.3 | (35.1, 53.9) |
| | | <50 | 8.6 | (5.6, 12.5) | 7.6 | (2.5, 16.8) | 0 | (0, 30.8) |
| | | 50-69 | 31.4 | (28.6, 34.3) | 30.7 | (24.7, 37.3) | 26.7 | (12.3, 45.9) |
| | | 70-79 | 56.5 | (53.2, 59.6) | 59.4 | (52.5, 65.9) | 42.9 | (27.7, 59) |
| | | 80+ | 66.5 | (63.6, 69.3) | 67.8 | (61.9, 73.2) | 75.8 | (57.7, 88.9) |
| | **Oxygen only** | **Overall** | 35.1 | (34.6, 35.7) | 32.5 | (31.4, 33.6) | 21.3 | (18.9, 23.7) |
| | | <50 | 3.6 | (3, 4.4) | 3.1 | (1.8, 5) | 2.6 | (0.7, 6.5) |
| | | 50-69 | 15.2 | (14.3, 16) | 15.2 | (13.4, 17.2) | 9.2 | (6.1, 13.3) |
| | | 70-79 | 38.1 | (36.9, 39.3) | 32.3 | (30, 34.7) | 24.2 | (19.1, 29.8) |
| | | 80+ | 52.7 | (51.8, 53.6) | 44.6 | (42.9, 46.3) | 33.2 | (28.9, 37.7) |

Table E11

**A**: Natural effects mediation model for respiratory support/ICU ward. Exposure: week admission (continuous). Confounders: age, sex, deprivation, comorbidity, severity (respiratory rate, oxygen saturations, GCS, serum urea, serum CRP). Mediators: respiratory support*icu/ward. Exposure-mediator interaction included. 10 imputed datasets used, and results combined using Rubin's rules.

| | estimate | standard error | Z | P | exp (estimate) | L95 | U95 |
|---|---|---|---|---|---|---|---|
| Pure natural direct effect | -0.167 | 0.014 | -12.280 | <0.001 | 0.847 | 0.824 | 0.869 |
| Total natural direct effect | -0.183 | 0.013 | -14.487 | <0.001 | 0.833 | 0.812 | 0.853 |
| Pure natural indirect effect | -0.043 | 0.005 | -9.581 | <0.001 | 0.958 | 0.949 | 0.966 |
| Total natural indirect effect | -0.060 | 0.003 | -19.587 | <0.001 | 0.942 | 0.936 | 0.947 |
| Total effect | -0.226 | 0.014 | -16.215 | <0.001 | 0.797 | 0.776 | 0.819 |

Approximate proportion mediated on risk difference scale = 24.3%.

**B**: Natural effects mediation model steroid use. Exposure: week admission (continuous). Confounders: age, sex, deprivation, comorbidity, severity (respiratory rate, oxygen saturations, GCS, serum urea, serum CRP). Mediators: respiratory support*icu/ward. Exposure-mediator interaction included. 10 imputed datasets used, and results combined using Rubin's rules.

| | estimate | standard error | Z | P | exp (estimate) | L95 | U95 |
|---|---|---|---|---|---|---|---|
| Pure natural direct effect | -0.289 | 0.013 | -22.583 | <0.001 | 0.749 | 0.731 | 0.768 |
| Total natural direct effect | -0.285 | 0.013 | -22.756 | <0.001 | 0.752 | 0.733 | 0.77 |
| Pure natural indirect effect | 0.005 | 0.002 | 2.641 | 0.008 | 1.005 | 1.001 | 1.008 |
| Total natural indirect effect | 0.008 | 0.001 | 6.606 | <0.001 | 1.008 | 1.006 | 1.011 |
| Total effect | -0.281 | 0.013 | -21.842 | <0.001 | 0.755 | 0.736 | 0.774 |

Approximate proportion mediated on risk difference scale = -2.5%

**C**: Natural effects mediation joint model for respiratory support/ICU ward and steroid use. Exposure: week admission (continuous). Confounders: age, sex, deprivation, comorbidity, severity (respiratory rate, oxygen saturations, GCS, serum urea, serum CRP). Mediators: respiratory support*icu/ward*steroid use. Exposure-mediator interaction included. 10 imputed datasets used, and results combined using Rubin's rules.

| | estimate | standard error | Z | P | exp (estimate) | L95 | U95 |
|---|---|---|---|---|---|---|---|
| Pure natural direct effect | -0.173 | 0.015 | -11.743 | <0.001 | 0.841 | 0.817 | 0.866 |
| Total natural direct effect | -0.189 | 0.013 | -14.280 | <0.001 | 0.827 | 0.806 | 0.849 |
| Pure natural indirect effect | -0.040 | 0.005 | -7.398 | <0.001 | 0.961 | 0.951 | 0.971 |
| Total natural indirect effect | -0.056 | 0.004 | -15.877 | <0.001 | 0.946 | 0.939 | 0.952 |
| Total effect | -0.229 | 0.015 | -15.002 | <0.001 | 0.795 | 0.772 | 0.82 |

Approximate proportion mediated on risk difference scale = 22.2%.

### Figures

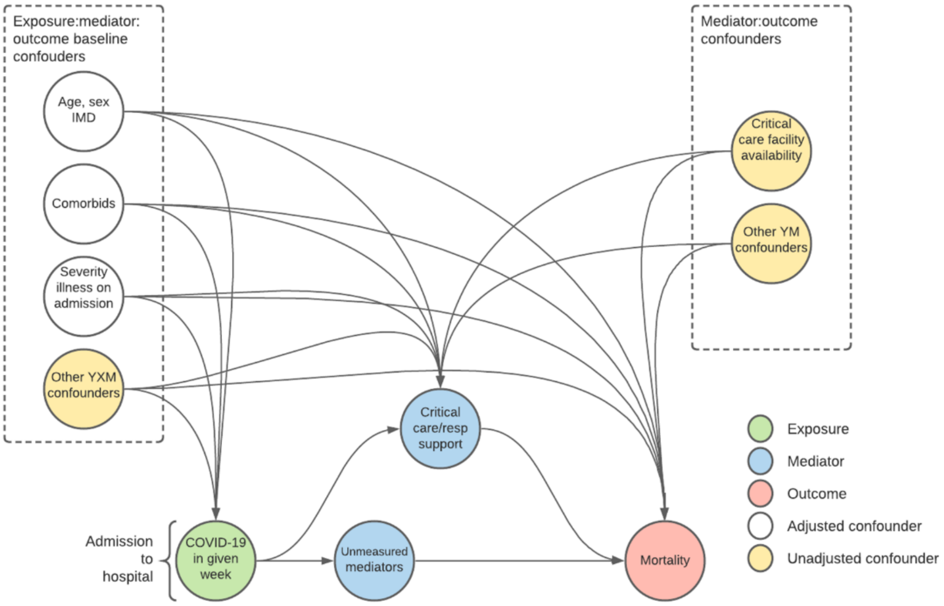


Figure E1: Directed acyclic graph showing putative causal model for analysis.

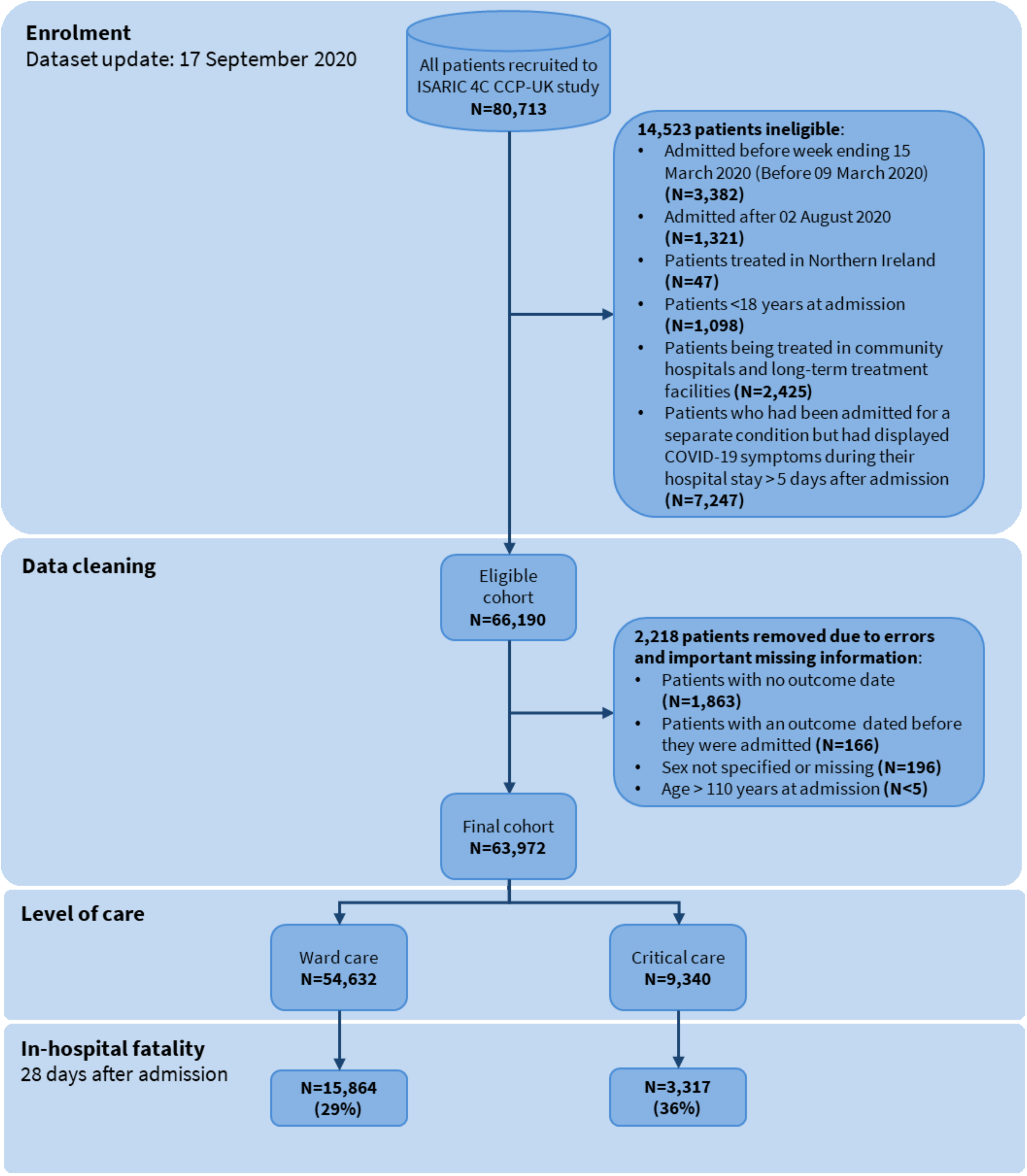


Figure E2: Consort diagram

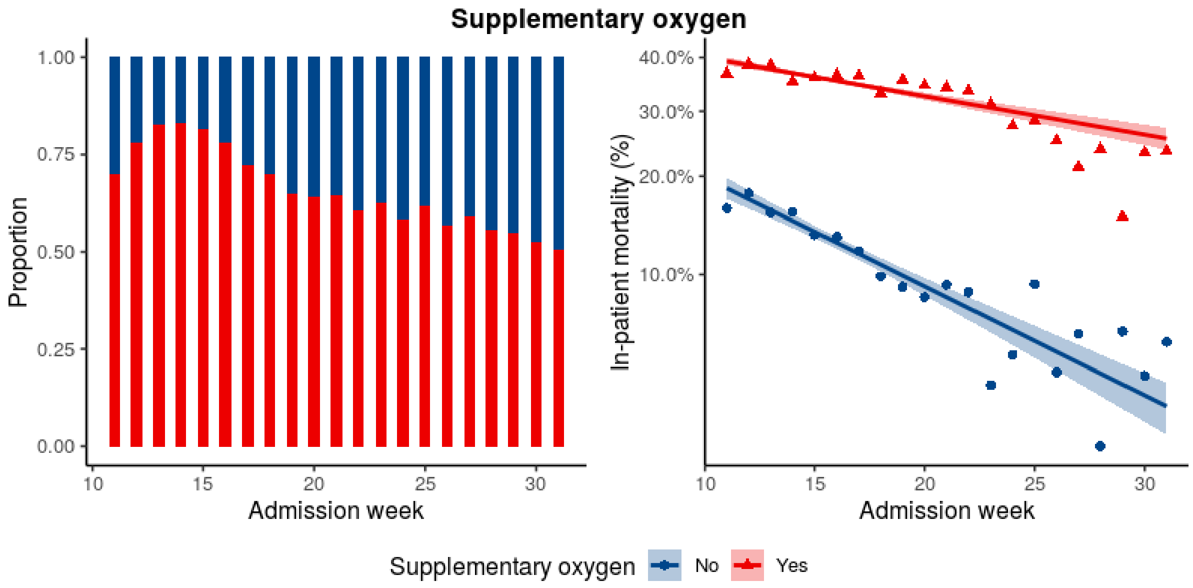


Figure E3: (Left) Changes in the proportions of all patients who received supplementary oxygen over time. (Right) In-hospital mortality rate per category over time

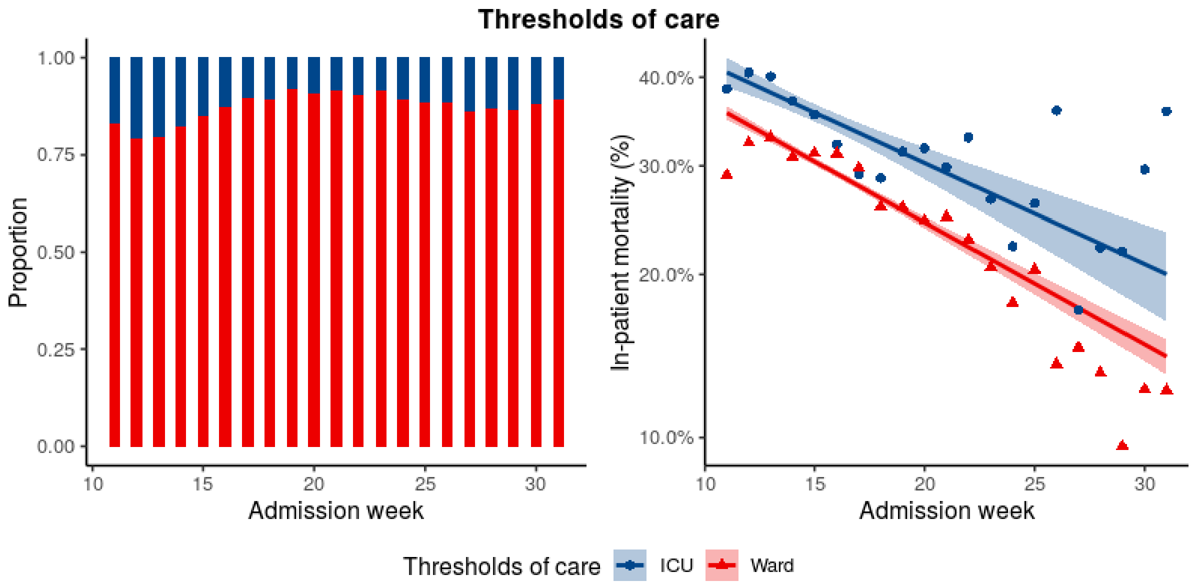


Figure E4: (Left) Changes in the proportions of threshold of care (ICU/Ward) over time. (Right) In-hospital mortality rate per category over time

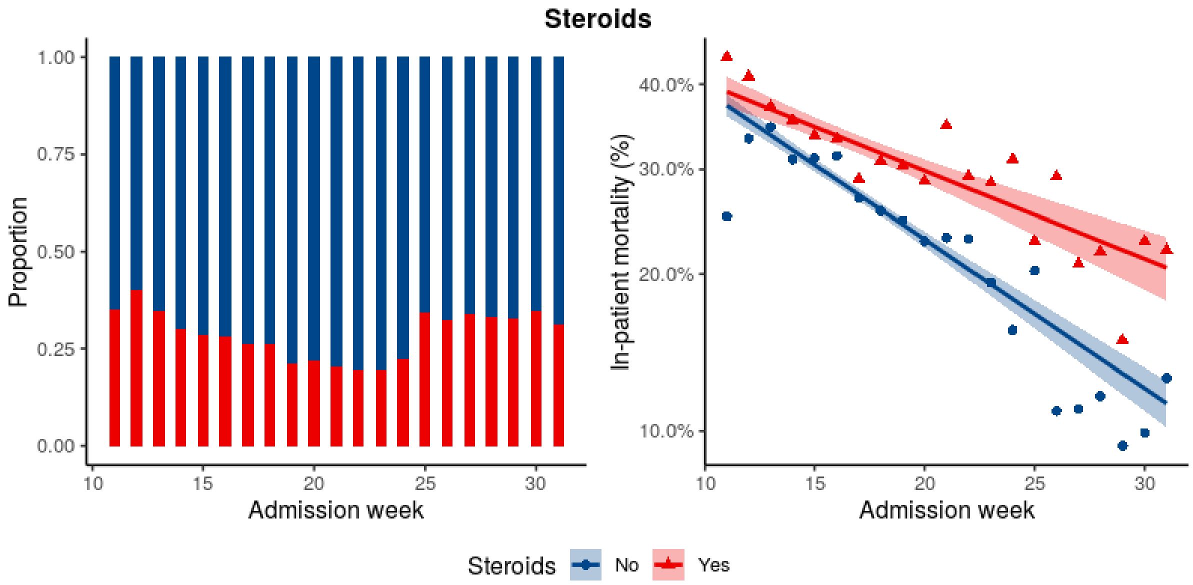


Figure E5: (Left) Changes in the proportions of all patients who received steroids over time. (Right) In-hospital mortality rate per category over time. Unknown measurements are excluded from this figure.

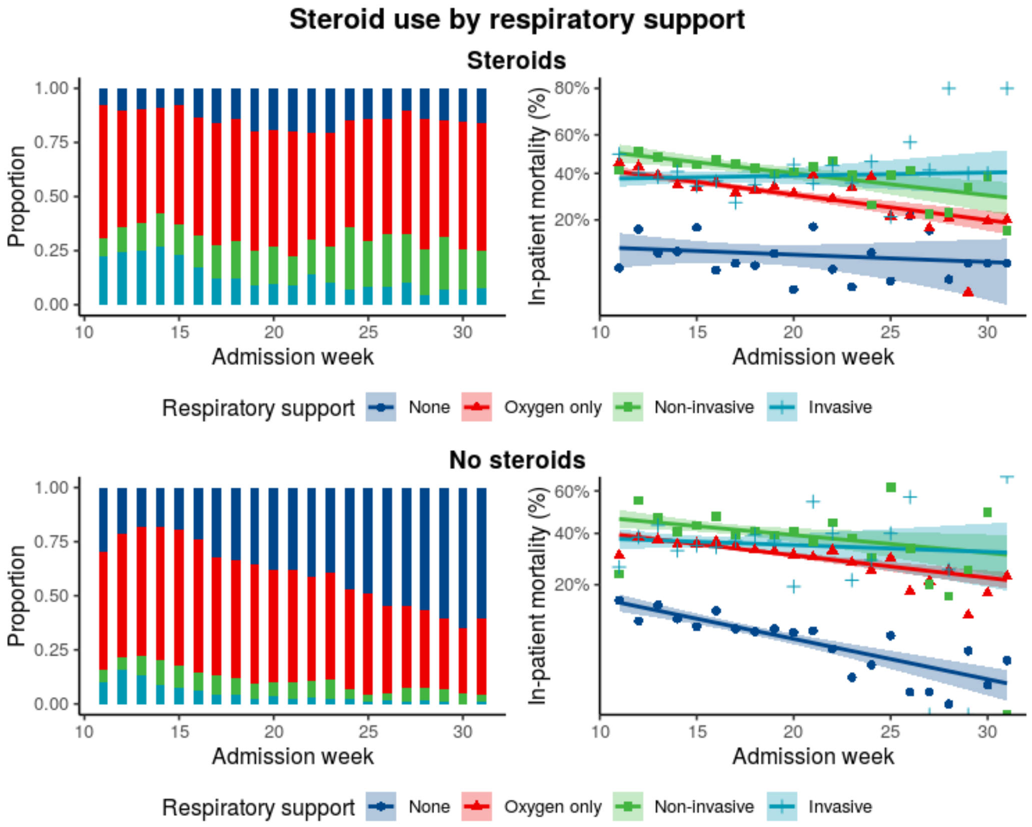

Figure E6: Respiratory support within steroid users (top) and non-steroid users (bottom). (Left) Proportion of respiratory support treatments by week of admission. (Right) In-hospital mortality rate per category by week of admission.

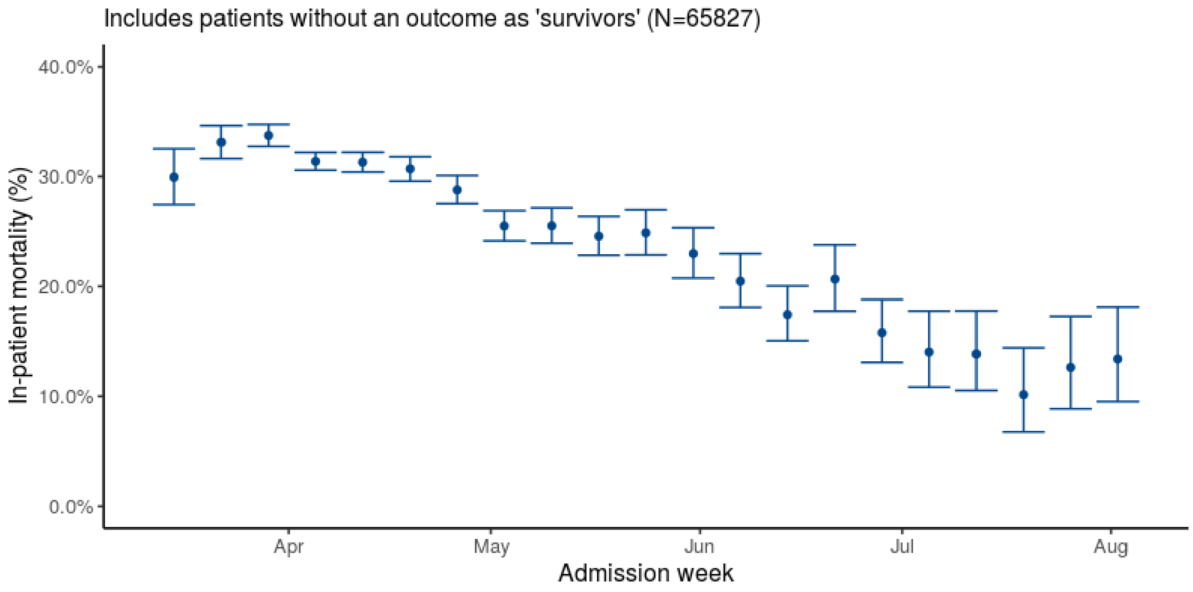


Figure E7: Sensitivity analysis - unadjusted weekly in-hospital mortality and 95% confidence intervals for patients admitted with SARS-CoV-2 from 9 March 2020 to 2 August 2020 Includes those without an outcome dated as ‘survivors’. Divided into 3 equal time periods (Weeks 11 to 17, 18 to 24 and 25 to 31). Confidence intervals calculated via exact method.
